# “Alexa, I just ate a donut”: A pilot study collecting food and drink intake data with voice input

**DOI:** 10.1101/2022.06.28.22276999

**Authors:** Louise AC Millard, Laura Johnson, Samuel R Neaves, Peter A Flach, Kate Tilling, Deborah A Lawlor

## Abstract

**Background:** Voice-based systems such as Amazon Alexa may be useful to collect self-reported information in realtime from participants of epidemiology studies, using verbal input. We demonstrate the technical feasibility of using Alexa, investigate participant acceptability, and provide an initial evaluation of the validity of the collected data. We use food and drink information as an exemplar.

**Methods:** We recruited 45 staff and students at the University of Bristol (UK). Participants were asked to tell Alexa what they ate or drank for 7 days, and also to submit this information using a web form. Questionnaires asked for basic demographic information and about their experience during the study and acceptability of using Alexa.

**Results:** Of the 37 participants with valid data, most were 20-39 years old (N=30; 81%) and 23 (62%) were female. Across 29 participants with Alexa and web entries corresponding to the same intake event, 357 Alexa entries (61%) contained the same food/drink information as the corresponding web entry. Participants often reported that Alexa interjected, and this was worse when entering the food and drink information compared with the event date and time. The majority said they would be happy to use a voice-controlled system for future research.

**Conclusions:** While usability of our skill was poor, largely due to the conversational nature and because Alexa interjected if there was a pause in speech, participants were mostly open to participating in future research studies using Alexa. Many more studies are needed, in particular, to trial less conversational interfaces.

**KEY MESSAGES:**

- Over the last few years voice-controlled ‘smart’ systems have emerged giving the possibility of collecting self-reported data using a voice-based approach.
- We successfully collected epidemiology food and drink information in real-time, demonstrating that voice-based collection of self-reported data is technically feasible.
- The conversational design of our skill meant that usability was poor, for example, most participants (86%) reported that Alexa either occasionally, often or always interjected during use, and the majority of participants who had previously used a paper diary or my fitness pal did not find Alexa as efficient to use compared with these approaches.
- After participating in this study, the majority of participants would be happy to use Alexa again, either at home or on a wearable device.
- Our results highlight that further work is needed to evaluate use of voice-based systems, including comparing Amazon Alexa with the Google Assistant, and trialling less conversational interfaces.

## INTRODUCTION

Epidemiology cohorts typically collect data at widely spaced time points (e.g. every 1-5 years)^1,2^. While some types of traits (e.g. weight or height) are fairly stable or change gradually across time, others such as activity levels, blood glucose levels, mental wellbeing, and dietary intake can vary more acutely, for example, within days, hours or even minutes. For these traits, prospectively capturing how they vary across time means we can assess how this variability relates to other traits and disease. Some acutely varying traits can be collected continuously and objectively using wearable digital devices, for example physical activity using accelerometers or blood glucose using continuous glucose monitors^3^. For others, such as mental health traits and dietary intake, no objective approach to measuring within-day variation in these traits exists, and they need to be collected via self-report.

One possible approach to providing real-time self-reported information is verbal input, which could enable participants to conveniently enter free-text rather than selecting from a set of prescribed options (in contrast to Ecological Momentary Assessment (EMA) ^4–6^). Over the last few years voice-controlled ‘smart’ systems have been released by several technology companies. These systems, such as Amazon Alexa and the Google Assistant, allow users to talk to a device rather than typing or pressing a button. They each have core functionality available by default (e.g. saying the time when asked for this), but also have developer platforms that allow anyone to produce and publish a custom voice-based app. This means it is now technically possible to collect self-reported data continuously across a day or several days, using verbal input.

Voice-based data collection may be most useful (in contrast to EMA) for collecting self-reported data that is both complex and variable across a day. One possible example is the food and drink a person consumes, and when they consume it. Traditionally, cohorts have collected dietary intake information using paper or online food frequency questionnaires or (less commonly) diaries. Limitations of these include retrospective recording, needing converting to electronic form, potential for missing data because participants are not prompted for missing information, and the inconvenience of having to carry a diary. More recently, some other approaches have been developed; a web-based dietary recall tool^7^ and an approach using photographs^8^. While these methods are able to collect detailed dietary information, they are burdensome so can only be used for short periods by highly motivated participants^9^.

In this pilot study we explore the potential of voice-based data collection in epidemiology research, using food and drink diaries as an exemplar. We have three key aims: 1) to demonstrate the technical feasibility of collecting data using Alexa, 2) to gain an initial insight into participant acceptability, and 3) to provide an initial evaluation of the validity of the collected data. In general, we view the capture and processing of the information as separate steps, and in this study focus on demonstrating and evaluating the former.

## METHODS

Ethical approval was given by the University of Bristol Faculty of Health Sciences Research Ethics Committee (approval number 63861).

### Study participants

Power calculations based on two measures suggested a sample size of at least 35 is needed (see details in Supplementary section S1). We recruited volunteers from University of Bristol staff and postgraduate student email lists. Participants were compensated with a £30 voucher.

### Description of system architecture: a voice-based system using Amazon Alexa

In this work we use the Amazon Alexa voice system. The Alexa system enables development of custom functionality, referred to as a custom skill. Alexa skills are comprised of intents that each define an interaction that a user can have with the skill. We developed a custom skill to collect food and drink intake events, with intents that allow participants to: 1) Add the date and time of an intake event, 2) add one or more items they ate or drank at this time, 3) cancel the event, 4) cancel the last item added to the event, and 5) submit the event. See example utterances in Supplementary table S1 and an example conversation in Figure 1. Supplementary section S2 provides further details.

**Figure 1:**
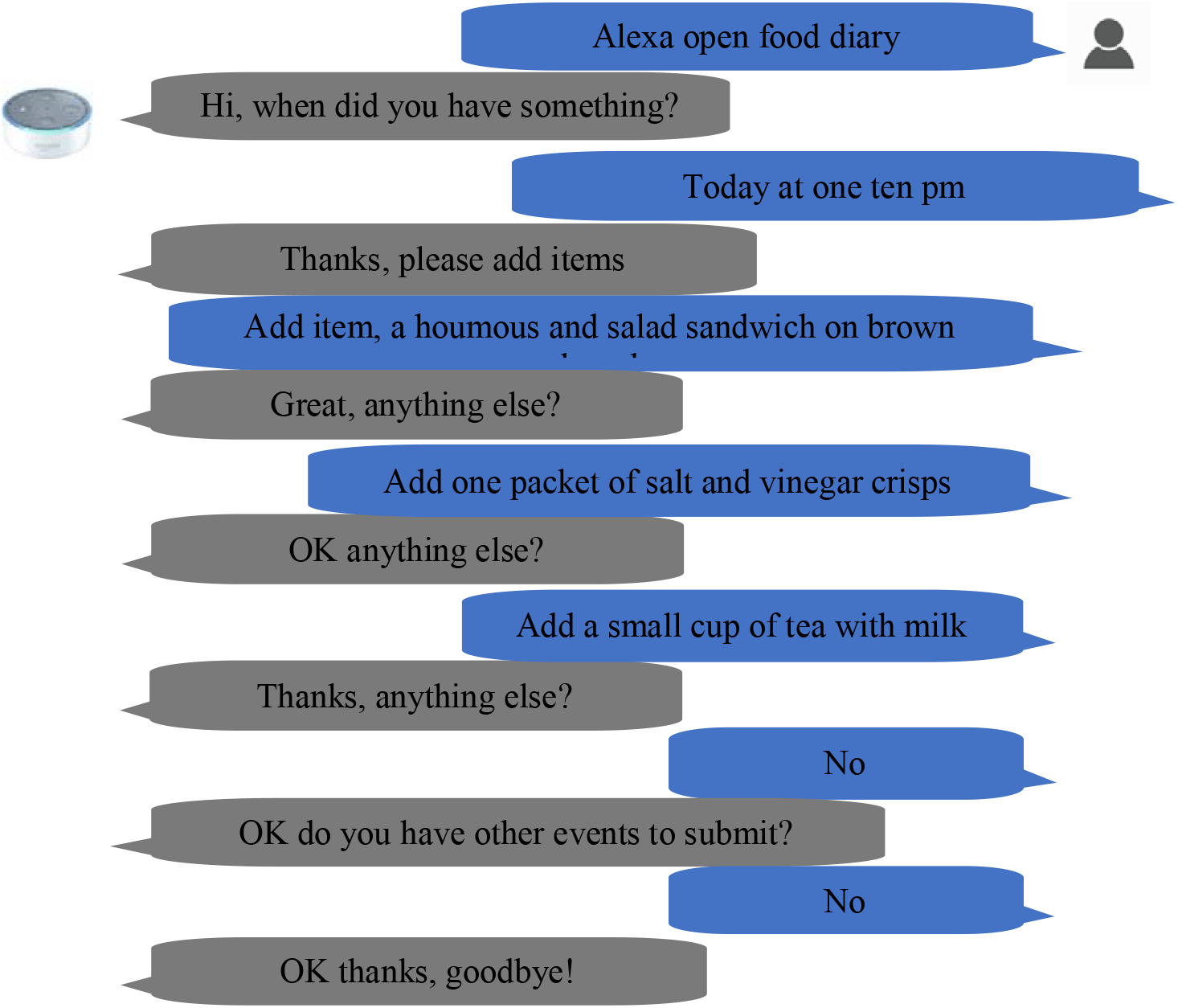
Example conversation of study participant with custom food diary skill

### Data collection protocol

An overview of the data collection protocol is shown in Figure 2. Due to the COVID-19 pandemic, participants took part at home. An initial email with accompanying participation information sheet (Supplementary file 1) invited staff and students to take part in this study. On replying, participants were sent a pre-participation questionnaire asking for basic demographic information such as their age and sex (Q1, Supplementary file S2). On completion, participants were booked for a 7-day Alexa data collection period.

**Figure 2:**
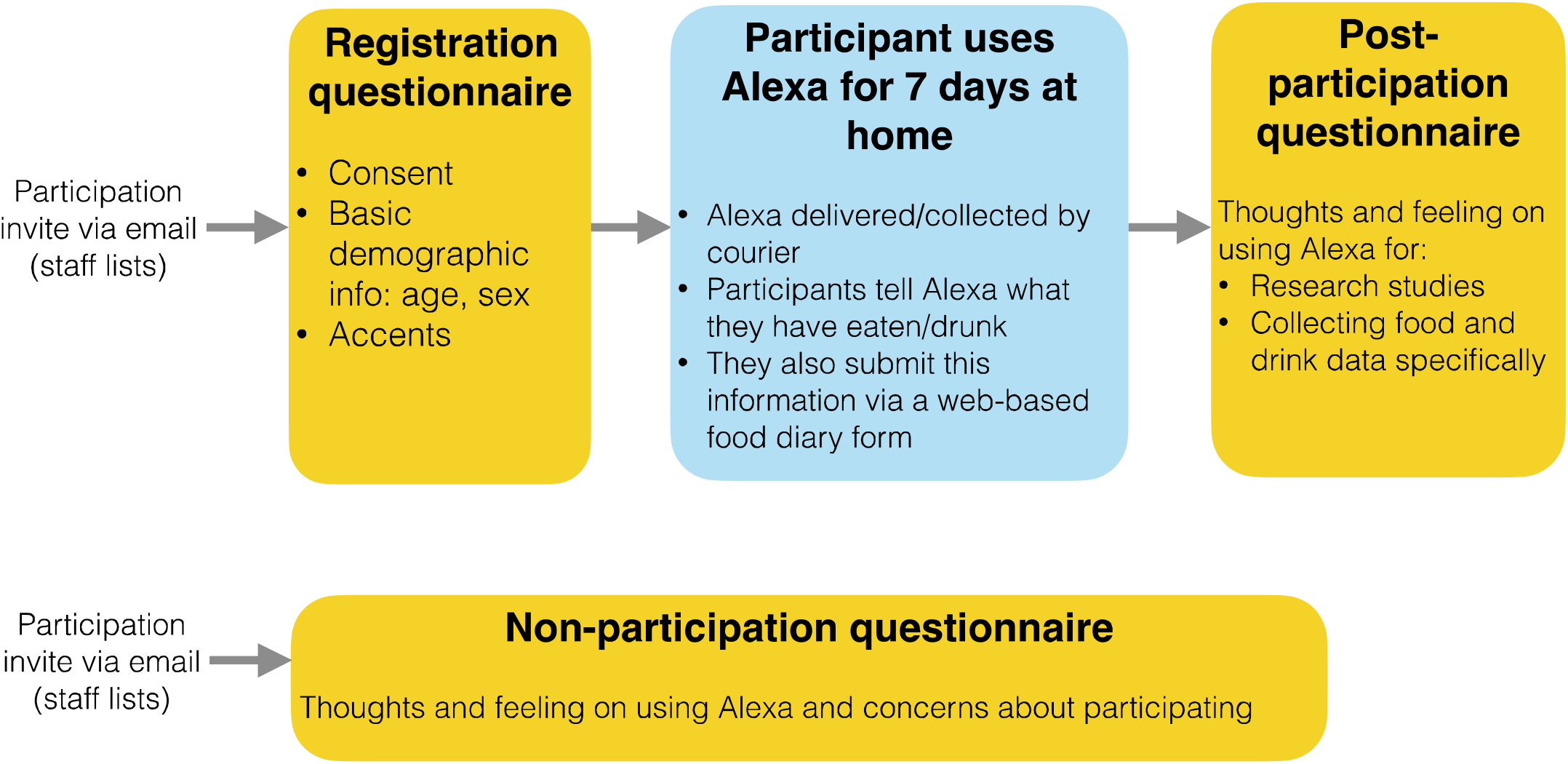
Overview of data collection protocol

The equipment was kept at the home of the principal investigator (LACM). On day 1 of a participants’ Alexa data collection period the equipment was delivered to their home by courier, along with a participant guide (Supplementary file 3). The participant was asked to set up the equipment and start using it as soon as they were able. Participants were instructed “After you have had something to eat or drink, we would like you to submit your food and drink information to Alexa <u>first</u>, and then submit it on the web form.” Entering the food and drink information using both Alexa and a web form (Q4, Supplementary file S2) allows us to compare the data entered using these approaches (i.e. relative validity^10^). On day 7 the equipment was returned to the principal investigator’s home by courier. Participants were then asked to complete a post-participation questionnaire on their experiences during the study and acceptability of using Alexa (Q2, Supplementary file S2).

To understand views on acceptability of using voice-based interfaces more widely (beyond our participant group) we also sent a further invitation (to the same email lists) asking for those who did not participate to complete a short questionnaire about their feelings on using voice-based devices and their reasons for not participating (Q3, Supplementary file S2).

Questionnaires were deployed via the University of Bristol REDCap secure web platform [https://sscmredcap.bris.ac.uk/redcap/]. Study emails are provided in Supplementary file S4.

### Analytical sample

A participant flow diagram is shown in Supplementary figure S1. Of the 45 participants who registered to participate, 1 withdrew, and 7 were excluded with incorrect equipment (see Supplementary section S3). The remaining 37 participants comprised our analytical sample. Of these, 1 participant did not attempt to use Alexa. In addition, 7 participants had Alexa entries, but no web diary entries completed within the 30 minutes directly following the Alexa submission. As Alexa and web entries must be within 30 minutes to be identified as corresponding to the same intake event in our data processing approach (see below), the Alexa and web entries from these participants could not be compared. The remaining 29 participants were used to compare the information entered via the web form versus Alexa (‘comparison’ sample).

### Data preprocessing

#### Mapping web food form entries to Alexa intake events

The web and Alexa entries both include the following information: a) intake timestamp: the date and time the participant (said they) ate or drank; b) submission timestamp: the date and time the participant submitted the entry; and c) intake items: one or more intake food and drink items. To compare the content of the web and Alexa entries, we first undertook an automated process to identify the Alexa and web entry pairs that correspond to the same intake event, referred to as counterpart entries. This was non-trivial because a participant might not have entered each entry with the web form immediately after entering it via Alexa, or the intake timestamp entered by Alexa might have been recorded incorrectly (i.e., Alexa might have heard the day or time stated by the participant incorrectly).

We identified counterpart entries using the intake and submission timestamps. The process we used was as follows (illustrated in Supplementary figure S5):

1) Identify counterparts as the set of entries where the web and Alexa intake timestamps were within 5 minutes of each other, and the Alexa submission timestamp is up to 30 minutes before the web submission timestamp. The non-exact match of the intake time was because participants can tell Alexa this using a phrase such as ‘just now’ or ‘ten minutes ago’ so may not correspond exactly to the intake time entered using the web form.
2) Identify web counterpart entries of the Alexa submissions not matched in step (1), as the nearest subsequent web entry where one occurs within 30 minutes of the Alexa entry.

#### Comparing counterpart web and Alexa entries food and drink description

We compared counterpart entries using two approaches, an automated approach, and a systematic manual approach.

##### Automated approach

We compared the text content of the counterpart entries, by comparing the set of words contained in each. Entries were preprocessed to remove plurality of words (e.g., crisps becomes crisp)^11^, and convert numbers to numeric values (e.g., “one” and “a” both become 1). For each counterpart pair we calculated the number of words in a) the web word set but not the Alexa word set, b) the Alexa word set but not the web word set, and c) both word sets.

##### Systematic manual approach

Our systematic manual approach was conducted by LACM. As this approach has some degree of judgement, we also asked 5 researchers independent to the project (within the same Unit but not involved in this study) to review 10 random entries (none repeated across researchers) so that we can evaluate inter-researcher variability of these manual evaluations.

To conduct this manual review we used a two-step process. First, the intake items of each counterpart pair were compared to determine if there was any similarity at all. If the set of items was completely different then these were marked as most likely corresponding to different intake events (i.e., the counterpart pairing did not work in this case – e.g. ‘a cup of coffee with milk’ versus ‘spaghetti bolognaise’). All other entries are taken forward to step two.

Step two involved reviewing each counterpart entry and for each, recording the number of food or drink items in a counterpart pair in the following categories:

a. Same item semantically (the two entries are equivalent with no additional or different information in each)
b. Same item but with different detail (e.g., ‘cup of tea’ vs ‘mug of tea’)
c. Same item, Alexa information has less detail (e.g., ‘cheese and salad sandwich’ vs ‘a sandwich’)
d. Same item, Alexa item has more detail
e. Same item, misspelling in Alexa input, but still understandable i.e., there is no loss of information (e.g. ‘to bagels’ versus ‘two bagels’)
f. Same item, misspelling in web form input, but still understandable
g. Same item, with Alexa entry issue, where the consumed item is still identifiable (e.g., “ball of yoghurt” rather than “bowl of yoghurt”)
h. Item with major entry issue, such that it contains no food or drink information, or the main essence of the meal/drink is missing (e.g. a “cough with milk” rather than “coffee with milk”)
i. Extra Alexa item with major entry issue (which can happen if a participant makes a mistake or then stops talking, then tries again so there is an extra item, e.g., “two”)
j. Extra Alexa item that is recognizable as a food or drink (i.e., should not be assigned to (i))
k. Extra web item

Supplementary table S2 shows some example assignments using this approach.

The independent researchers who completed 10 entries were provided with an information sheet describing the task (Supplementary file S5). We evaluated agreement between the assignments of LACM and the independent researchers visually using a stacked bar chart.

The automated and systematic manual approaches are complementary as the former is objective but is likely to be a more pessimistic assessment of agreement. This is because, participant may not write an entry the same way as they would speak it. For example, a participant might write “1 x apple. 1 bar of chocolate” but say “one apple and a chocolate bar”, which have differences in the words used even though they are semantically the same.

### Statistical analyses

#### Usage summary

We summarised the participants’ usage of the web and Alexa approaches using the median and interquartile range (IQR) of the number of submitted web and Alexa entries, respectively.

#### Comparison of counterpart diary entries

We compared the intake timestamps in the counterpart pairs using a plot similar a Bland-Altman plot but where the x-axis is the intake time entered using the web form rather than the average. Assuming the intake timestamp entered on the web form will be largely correct, this is to help show whether intake time submitted via Alexa may be less accurate for particular times of day. We summarised the automated and systematic manual comparisons using stacked bar charts.

#### Summarizing the number of incomplete attempts

The Alexa skill saves partial entries (i.e., those that have not been submitted perhaps because the internet connection was interrupted) in addition to completed entries. We estimated the median (IQR) number of unsuccessful attempts across participants.

#### Evaluating questionnaire responses on acceptability

We summarised responses of the participation questionnaire (Q2) and the non-participation questionnaire (Q3) by calculating the number of participants (and percentage) that responded to each questionnaire item option. Responses to free text items were read and reread in order to identify key themes.

The Alexa skill and web service code is available at https://github.com/MRCIEU/food-diary-alexa-pilot-aws/. Analysis code is available at https://github.com/MRCIEU/food-diary-alexa-pilot-analysis/ (git tag v0.1 corresponds to the version of the analyses presented here).

## RESULTS

### Participant characteristics

Participant characteristics are summarized in Table 1. Most participants were in early adulthood (81% 20-39 years old). Our sample included more female participants than male participants (62% female). The majority (84%) reported that they did not believe they had a strong UK regional accent, with 68% reporting they did not have an accent due to English being a second language. 43% of our participants have an Alexa at home that they use.

**Table 1:** Participant demographics

| **Participant characteristic** | **N (%)** |
| --- | --- |
| Age band – <20 | 4 (11%) |
| – 20-29 | 16 (43%) |
| – 30-39 | 14 (38%) |
| – 40-49 | 2 (5%) |
| – 50-59 | 1 (3%) |
| Sex – N female | 23 (62%) |
| Regional accent - No | 31 (84%) |
| - A little | 4 (11%) |
| - Yes | 2 (5%) |
| Non-English accent - No | 25 (68%) |
| - A little | 9 (24%) |
| - Yes | 3 (8%) |
| Has a voice-controlled device - No | 16 (43%) |
| - Yes but it's used by others, not me | 5 (14%) |
| - Yes and I use it | 16 (43%) |

These summaries use the usage sample, N = 37.

### Usage summary

On average, participants completed more web diary entries compared to Alexa entries (median number of entries 17 [IQR: 13, 27] compared with 11 [IQR: 7, 21]; paired t-test P value <0.001) (histograms shown in Supplementary figure S6). The median number of partial Alexa attempts was across all participants was 6 [IQR: 1, 9].

### Comparison of counterpart diary entries

#### Intake timestamp comparison of web form versus Alexa entry

Across all participants in the counterpart subsample 72% of the completed counterpart entries had a matching timestamp (Figure 3). The median proportion of completed counterpart entries with a matching timestamp across participants was 0.67 [IQR: 0.5, 1].

**Figure 3:**
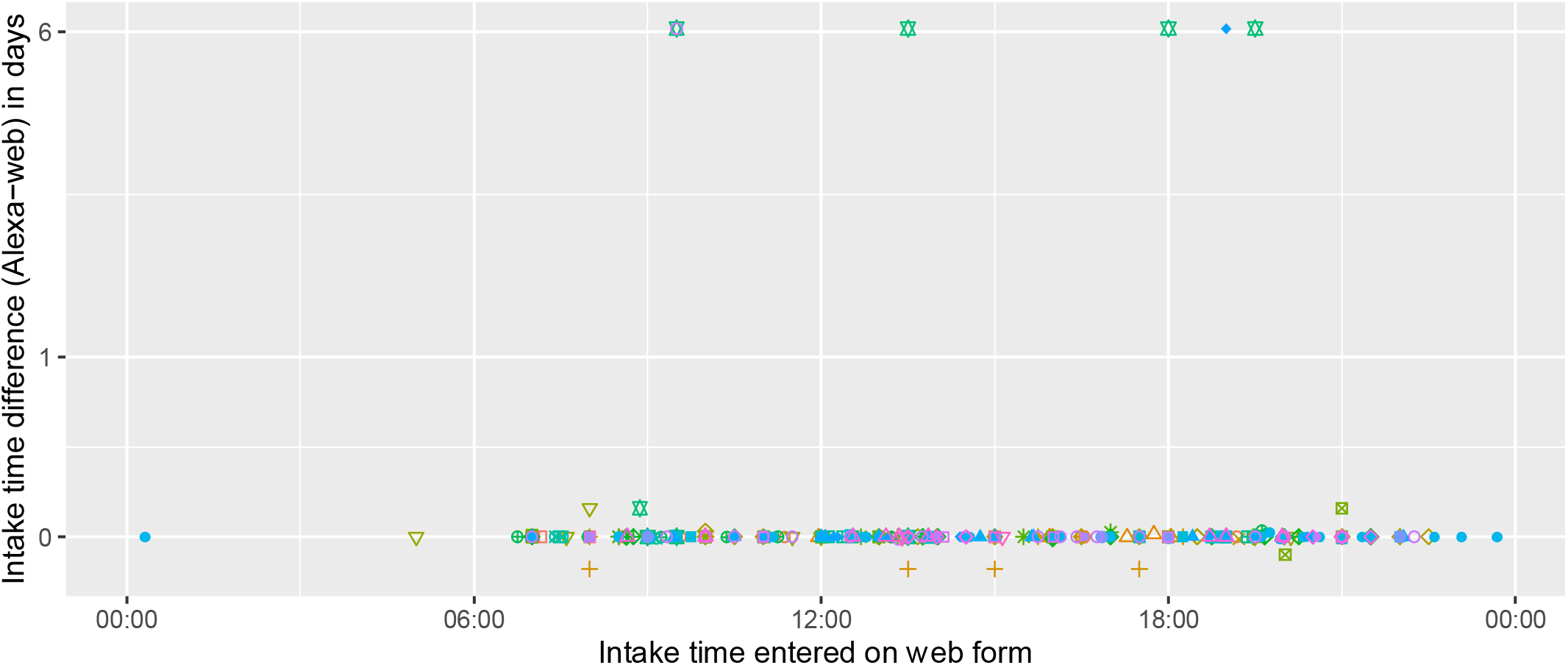
Plot comparing intake timestamps recorded using Alexa versus the web form Each colour/symbol combination denotes a particular participant. Each point refers to an intake event for a participant, and shows the intake event time entered using the web form (x-axis) and the difference between the intake event time entered using Alexa versus the web form (y-axis).

#### Food and drink description comparison of web form versus Alexa entry

Across the 29 participants in the comparison subsample there were 310 counterpart entries. Results comparing the submitted food and drink information using the automated and manual comparison approaches are shown in Figure 4 and Figure 5, respectively. Of the 310 counterpart entries manually reviewed, 21 (7%) were classified as corresponding to different intake events. The remaining 289 counterpart entries included 612 web form items and 588 Alexa items, with 33 extra web items (not identified in the counterpart Alexa entry) compared to 9 extra Alexa items (not found in the counterpart web entry). The majority (N=357 [58% and 61% for the web and Alexa items, respectively]) of the items entered via the web form and Alexa were the same, containing the same information. Of the 194 items that were identified as corresponding to the same intake item but containing different information, 64 (33%) had less detail from Alexa, 12 (6%) had more detail from Alexa, 15 (8%) had different detail, 3 (2%) items had a web entry issue, 36 (19%) had an Alexa entry issue, 4 items (2%) had spelling mistakes in the web version not the Alexa version, 59 (30%) had a misspelling in the Alexa only, and 1 (1%) had a misspelling in both the Alexa and web input. Of the 59 items with an Alexa misspelling, 40 of these (68%) were recording the word ‘to’ rather than ‘two’. 28 (5%) of the 588 items entered via Alexa were classified as having a major entry issue. We did not identify systematic differences in the assignments of LACM for the systematic manual approach compared to those of independent researchers (Supplementary figure S7).

**Figure 4:**
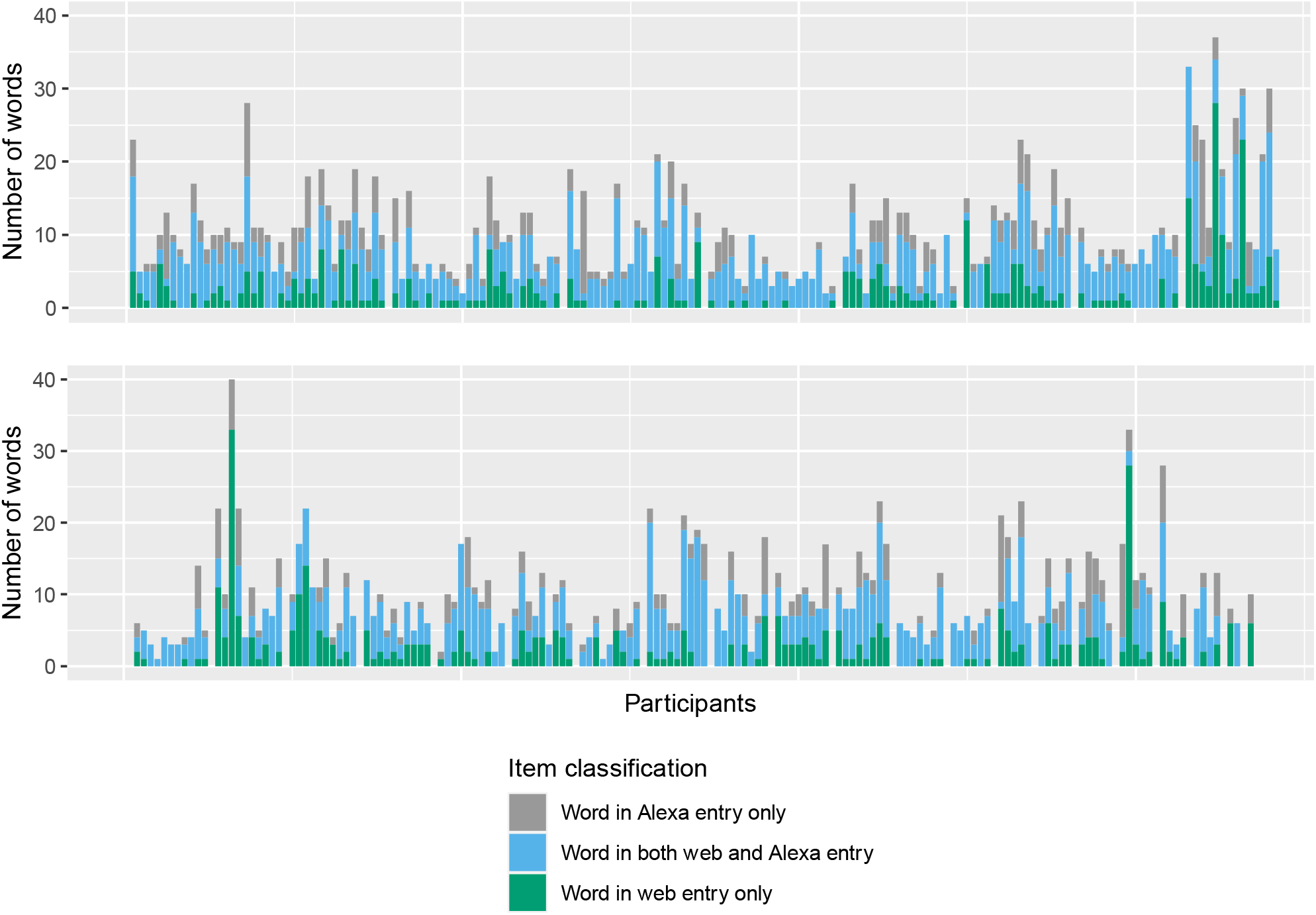
Summary of automated comparison of the web and Alexa submitted food and drink information Each stacked bar shows the number of unique words in a) both the web and Alexa entries, b) the Alexa entry only, and c) the web entry only. Each block of stacked bars shows the set of entries for a given participant.

**Figure 5:**
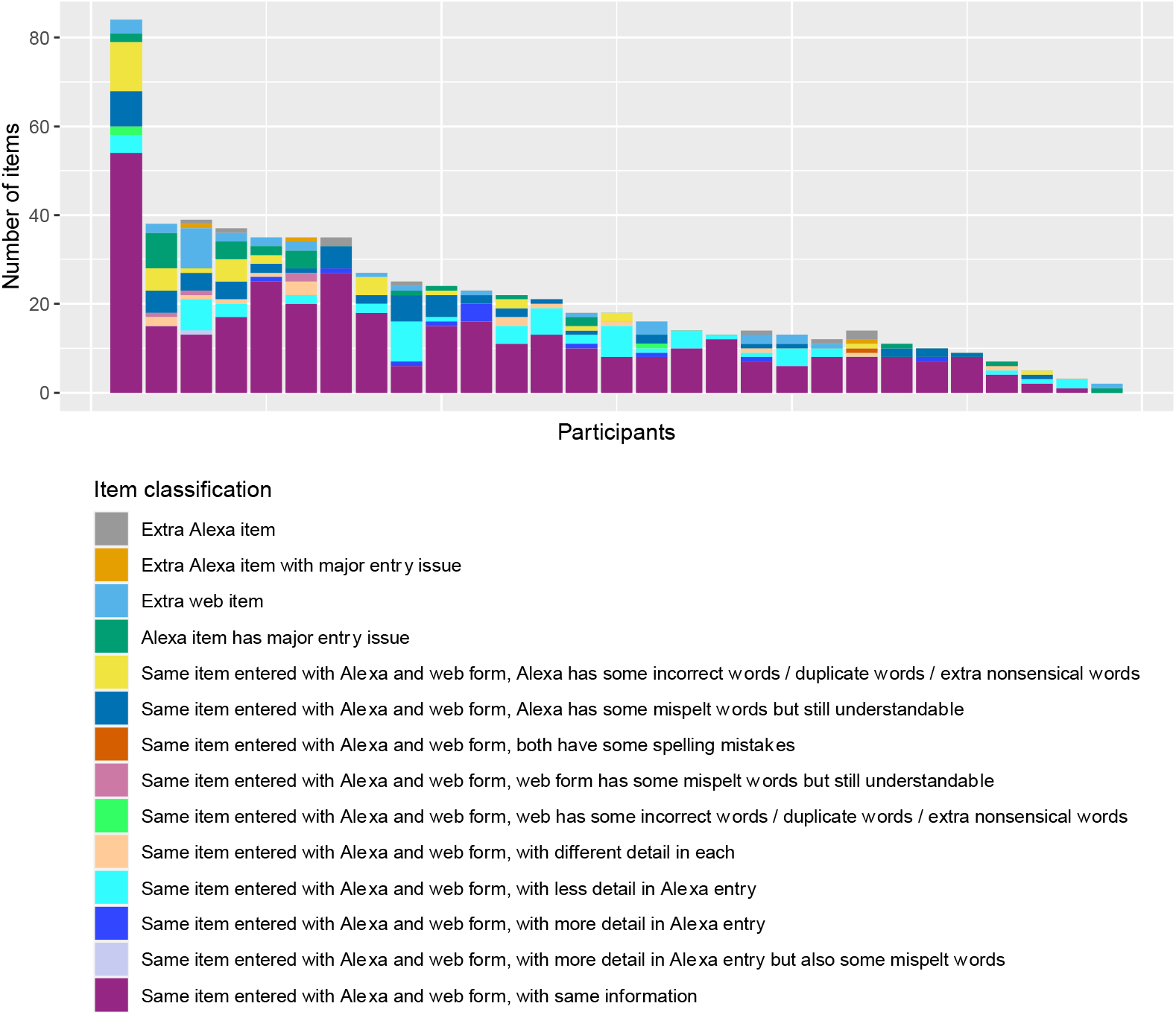
Summary of manual evaluation of the submitted food and drink information Each stacked bar shows the number of items in each category, for each participant. Manual evaluation conducted by LACM. Supplementary figure S7 shows comparison of assignments with those by independent assessors for random subsamples.

### Evaluating participants questionnaire responses on usability and acceptability

Summaries of post-participation questionnaire responses are provided in Table 2. Of the 35 participants who completed the post-participation questionnaire 26 (74%) said they would be happy to use voice-controlled system at home for future research and 28 (80%) said they would be happy to use one on a wearable device (e.g., a smart watch). Alexa sometimes interjected when participants were telling her when they ate or drank, with 7 participants (20%) saying this happened often or always, and 11 (31%) saying this happened occasionally. Alexa often interjected when participants were telling her what they ate or drank, with 18 participants (51%) saying this happened often or always and 12 (34%) saying this happened occasionally. In terms of convenience, enjoyment, and efficiency, 18 (51%), 21 (60%) and 15 (43%) participants, respectively, said they found using Alexa OK or better.

**Table 2:**
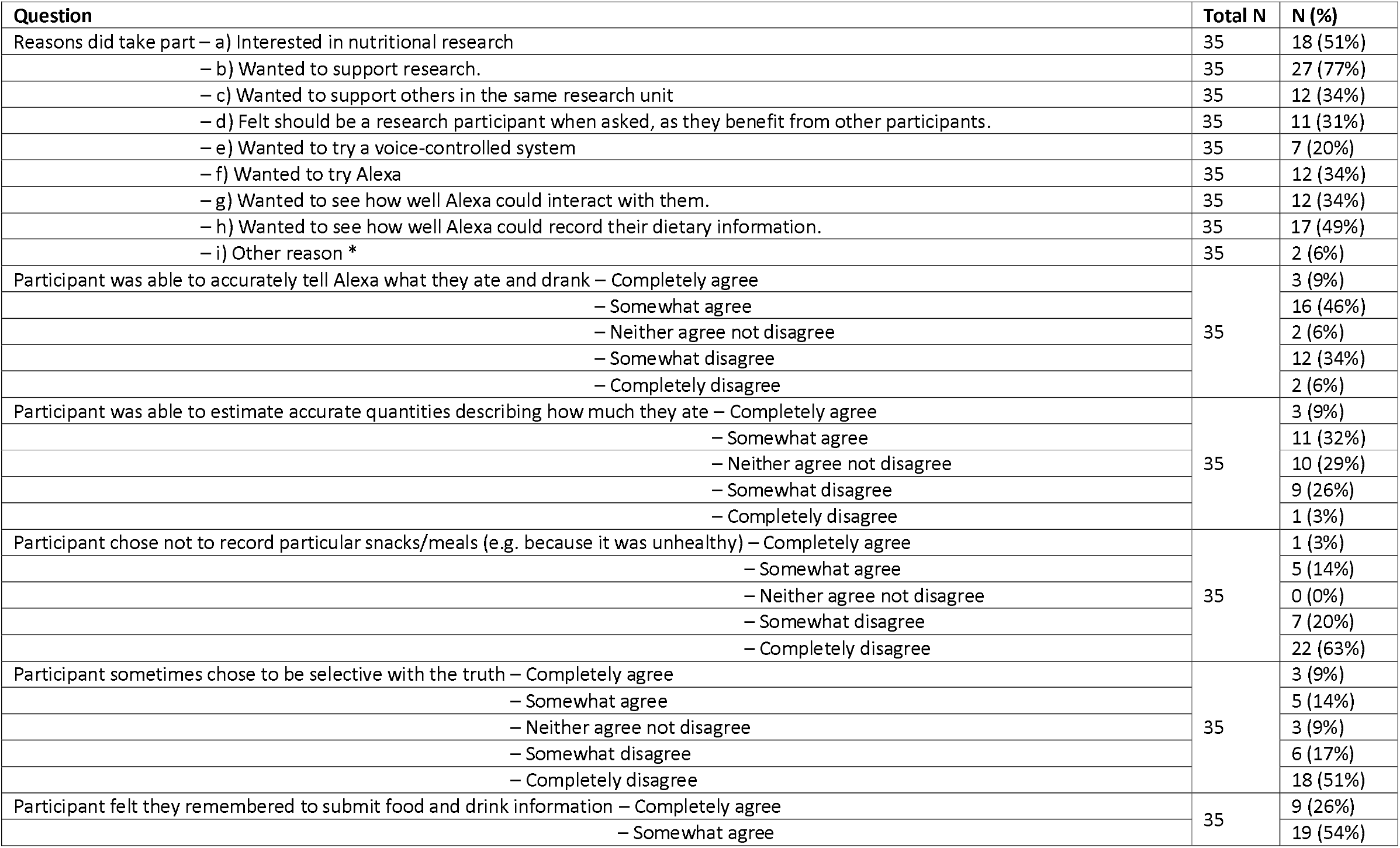
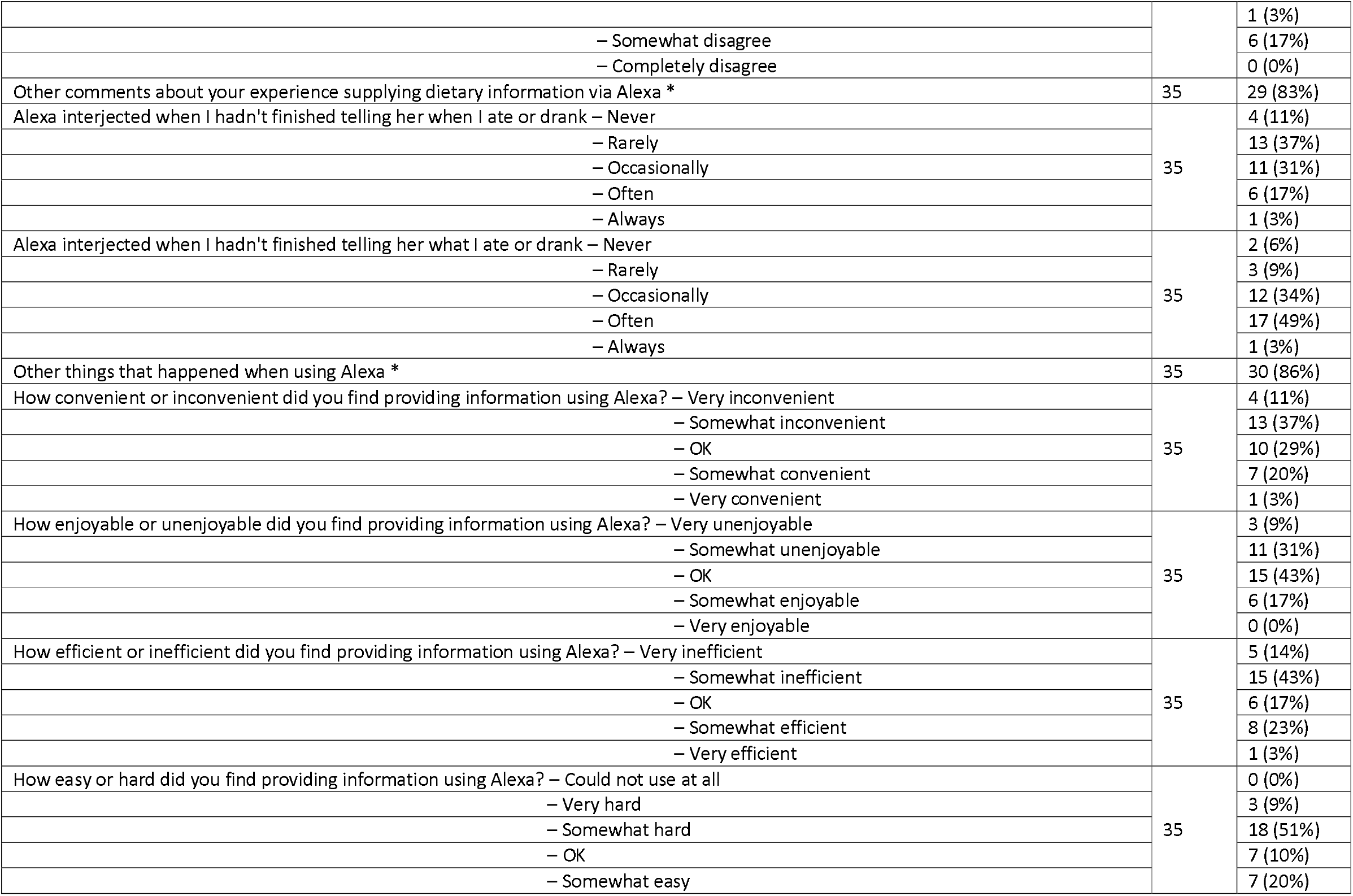
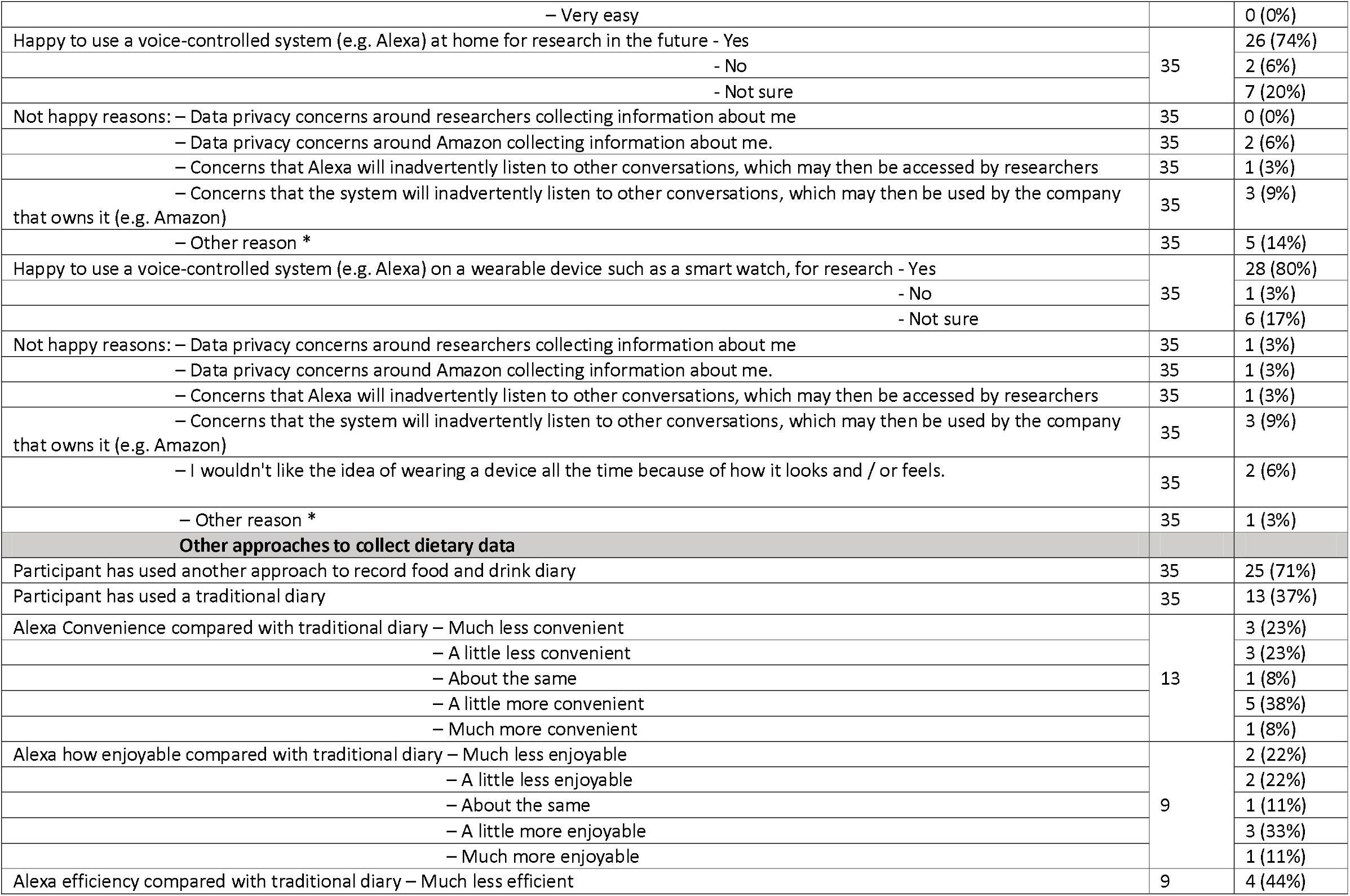
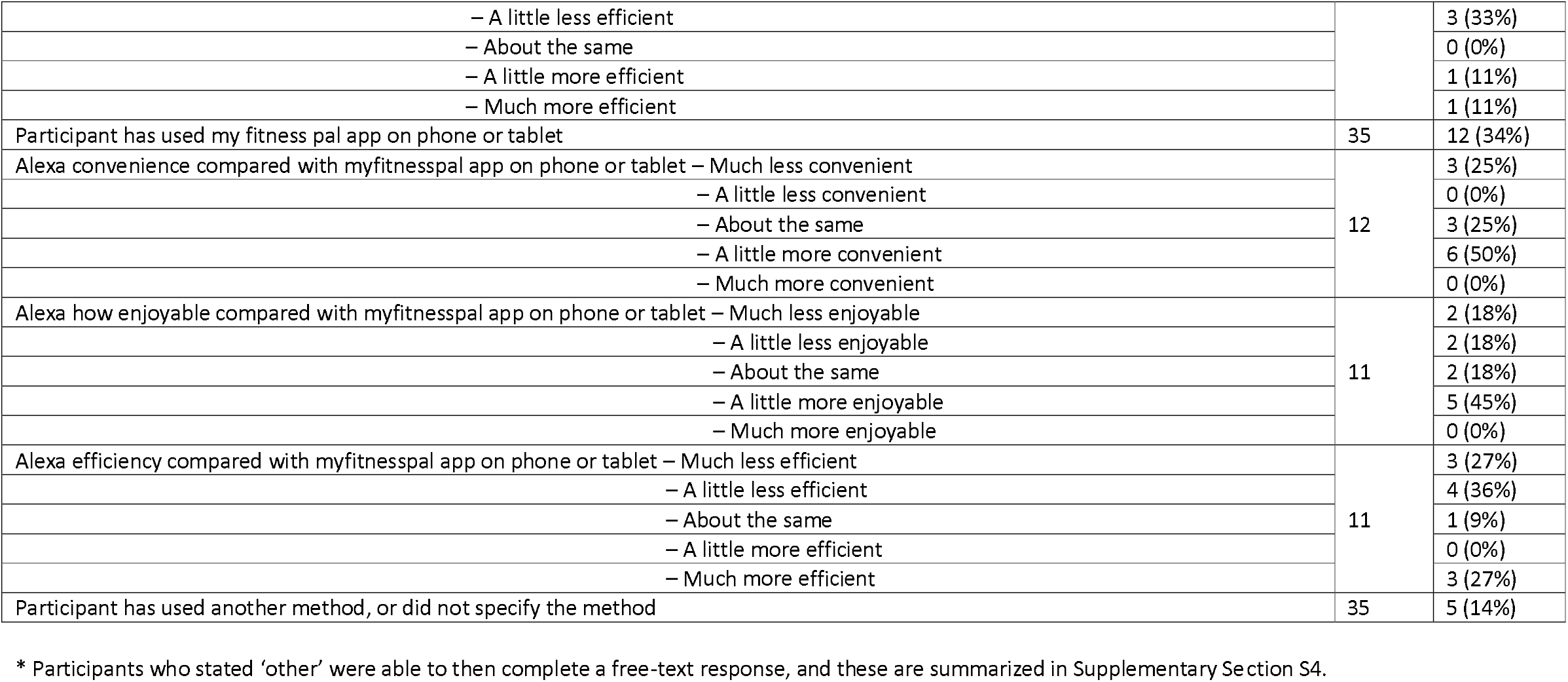
Post-participation questionnaire summary

Of the 35 participants who completed the post-participation questionnaire, 25 (71%) had previously used another approach to record their food and drink intake (Table 2). Of the 13 participants who have used a traditional diary (on paper or a computer), 7 (54%) found using Alexa at least as convenient, 5 (56%) at least as enjoyable, 2 (22%) at least as efficient. Of the 12 (34%) participants who have used MyFitnessPal, 9 (75%) found Alexa at least as convenient, 7 (64%) at least as enjoyable and 4 (36%) at least as efficient.

### Evaluating non-participation questionnaire responses

Of the 69 participants who responded, 11 (16%) did not take part due to privacy concerns (either around Amazon or researchers collecting their diet information or Alexa inadvertently listening to other conversations (Table 3). 61% (N=42) stated they would be happy to use Alexa at home for research in the future, while 57% (N=39) said they would be happy to use Alexa on a wearable device for research purposes.

**Table 3:**
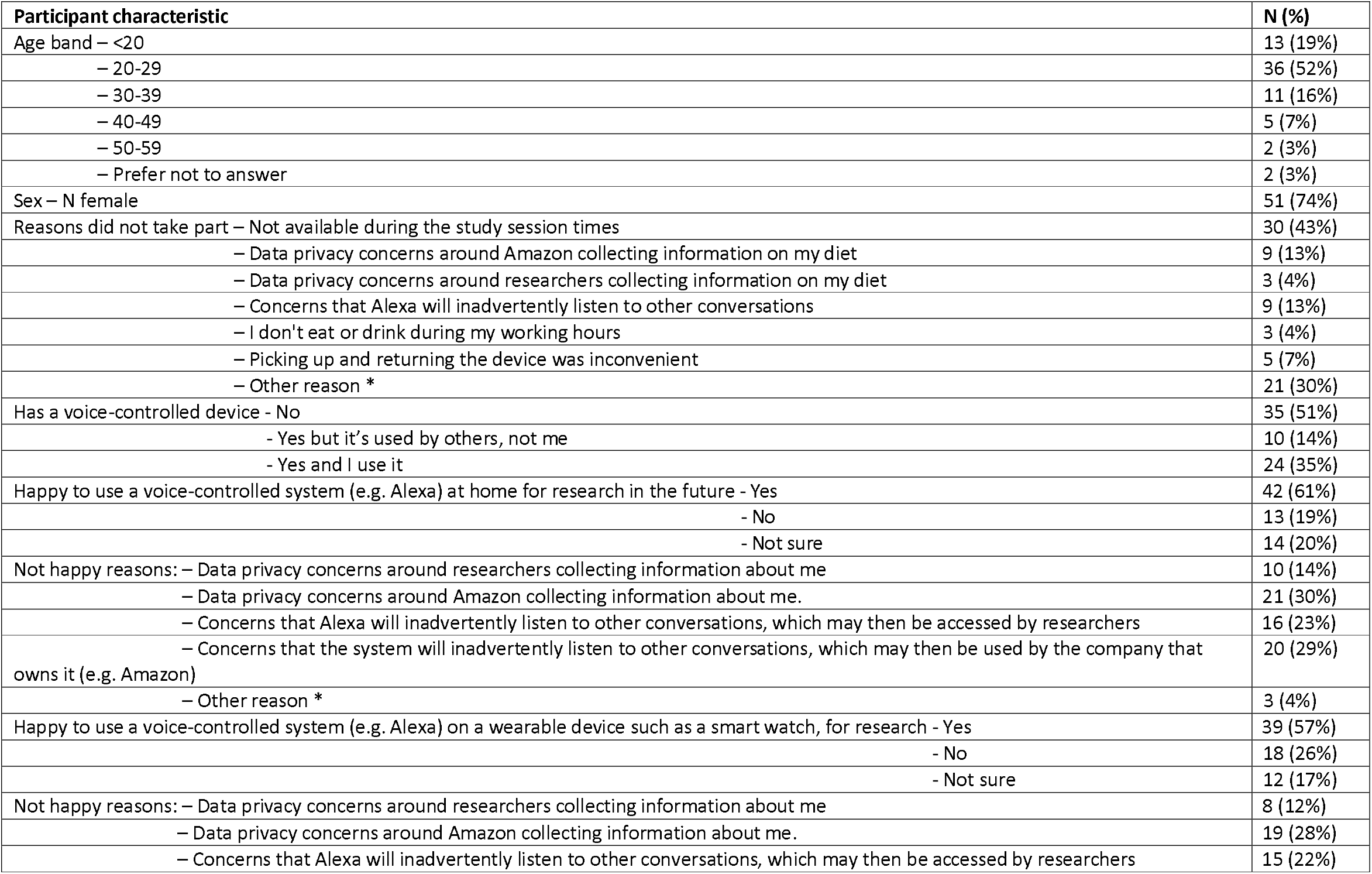
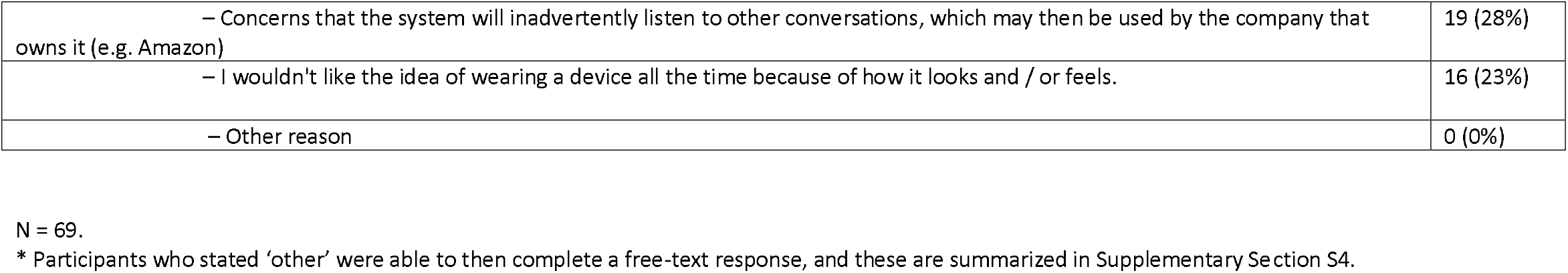
Non-participation questionnaire summary

## DISCUSSION

In this study we have demonstrated the technical feasibility of collecting data using Alexa for epidemiology research, by successfully developing an Alexa skill to collect food and drink information and using it to collect data in 37 participants across a period of 7 days (5 full days). Our results provide a useful initial insight into participant acceptability of using this approach and validity of the collected data. On average, more entries were submitted via the web form versus Alexa. Our results suggest that intake date and time was largely entered accurately via Alexa. The majority (61%) of Alexa entries contained the same food/drink information as the corresponding web entry, according to our systematic manual approach. The most common differences were the Alexa information having less detail or a homophone error (most often ‘to’ rather than ‘two’).

Overall, usability of our Alexa skill was fairly poor. The majority of participants reported that Alexa interjected while they were trying to enter food and drink information (34% sometimes, 51% often/always), with better results for the date/time of the intake event (31% sometimes, 20% often/always). Several participants reported finding it difficult to avoid pausing while articulating what they ate or drank, which might cause Alexa to interject or cut-out. Some reported reducing the information they provided so that Alexa would be more likely to accept it. Participants also reported that Alexa would sometimes not understand or exit the skill during use.

The voice interface we have trialed comprises of our Alexa skill implementation and the Amazon back-end logic, and only the former is under our control. The implementation and deployment of the Alexa skill has several components, with many choices regarding the design of the voice interface, the technical infrastructure, and the study protocol (e.g., location of data collection which was home-based in our study). Each of these may have affected the usability of the skill to collect food and drink information. Most notably, we conclude that the conversational interface of our skill (where participants first tell Alexa the time, then each of the items consumed) was not successful, as when the skill inadvertently cut out (e.g., because of multiple failed attempts conversing with Alexa or a poor internet connection) the participant would have to start that entry from the beginning. A less conversational interface where the participant states the information without the separate prompts, would likely be more usable. While our results suggest Alexa may be more appropriate for entering short summaries of information, in the longer term, integration of this approach with other approaches such as a phone app, can be used to supplement the voice-collected data. For example, using Alexa to log events directly after eating or drinking, then entering more detail via a phone app when convenient to do so.

Strengths of our study include the collection of pilot data ‘in the wild’ rather than in a controlled lab-based setting. We collected food and drink information via a web form in addition to Alexa, to allow comparison of the collected data via these approaches. Our study has several limitations. While asking participants to provide information via both Alexa and a web form was valuable, interactions with one of these approaches may have impacted their interaction and perceived feelings towards the other. The Alexa and web entries in our data had no explicit link and identifying entries that corresponded to the same intake event was difficult. We could only assess relative validity of the Alexa entries (relative to the web form entries) i.e., we have no absolute ground truth. While the intake date and time could be easily compared between the Alexa and web form entries in an automated way, comparing the free-text food and drink information was non-trivial as differences in the way the participant conveyed this information would not necessarily amount to meaningful differences in the submitted information. Most of these limitations could be rectified by integrating this voice-based approach with a phone app where the participant can review each entry and either correct it or mark it as correct, instead of requiring a web diary, so that validity can be assessed in an automated way by evaluating the corrections the participant makes. Further strengths and limitations and details are provided in Supplementary section S5.

While other studies have used voice-based approaches in other health settings^12,13^, to our knowledge, this is the first study to assess collecting self-reported epidemiology data with a voice-based system (a previous grant that sought to create a voice-based interface did not to our knowledge achieve this objective^14^). Many more studies are needed to understand the strengths and limitations of different approaches to collect epidemiology data using voice, for example, with different voice-based systems (e.g. comparing Amazon Alexa versus the Google Assistant), different voice interface designs particularly those that are less conversational, and to further evaluate biases in the collected data^15^. Voice-based approaches may be particularly useful in populations that might not be able to write (or write with ease), for example those with learning difficulties such as dyslexia, or certain diseases such as motor neurone disease.

## Data Availability

Data cannot be shared for privacy reasons.

## ETHICS APPROVAL

Ethical approval for this study was given by the University of Bristol Faculty of Health Sciences Research Ethics Committee (approval number 63861).

## AUTHOR CONTRIBUTIONS

LACM and SRN conceptualized the use of Alexa for epidemiology data collection. LACM conceptualized the study, led design of the study, wrote the first version of the manuscript. All authors contributed to the design of the study. All authors critically reviewed and revised the manuscript. LACM acts as the guarantor for this paper.

## DATA AVAILABILITY

Data cannot be shared for privacy reasons.

## SUPPLEMENTARY DATA

Supplementary data are available at IJE online

## ACKNOWLEDGEMENTS

We are extremely grateful to the participants that took part in this study. We are grateful to Mrs Shirley Jenkins for providing administrative support for this study. Thank you to Professor Katrina Turner for helpful feedback on the qualitative review of questionnaire responses. Thank you to Mr Tom Clark, Dr Emily Kawabata, Dr Michael Lawton, Dr Nancy McBride, and Dr Tom Palmer who conducted independent manual assessments of subsamples of the food and drink entries.

