## Supplementary information for "“Alexa, I just ate a donut”: A pilot study collecting food and drink intake data with voice input"

Millard et al.

SUPPLEMENTARY MATERIAL

### SUPPLEMENTARY TEXT

#### Supplementary section S1: Power calculations to determine appropriate sample size

As described in the introduction we aimed to assess both technological feasibility and participant acceptability. We defined primary measures of each of these on which to base our power calculations.

Acceptability
Primary measure: the proportion of participants that said they would be happy to use Alexa for research in the future (for both home and body-worn devices).
Power calculation statistic: Precision of the primary measure
Anticipated true proportion: 0.9
Desired precision (half width of CI): 0.1 (i.e. CI: 0.80-0.90]
Confidence level: 0.95
**Sample size required: 35**
*Calculated using: https://epitools.ausvet.com.au/oneproportion*

Feasibility
Primary measure of feasibility: the average accuracy of submitted food and drink data compared to web form (i.e. the proportion of intake events containing the same information).
Power calculation statistic: precision of the primary measure.
Anticipated true average accuracy: 0.8
Desired precision (half width of CI): 0.05
Sample size required: **22**
*Calculated with Stata code: ciwidth onemean, probwidth(0.8) width(0.1) sd(0.1)*

#### Supplementary section S2: Technical details of system architecture

In this study we use the Amazon Alexa voice system as it is used most widely and has the most mature developer capabilities (in comparison with the Google Assistant).

Custom skills are set up using the Alexa Skills Kit, which comprises of the Alexa developer console, an application programming interface (API), documentation and code samples [https://developer.amazon.com/en-US/alexa/alexa-skills-kit]. The general infrastructure required for an Alexa skill is shown in Supplementary figure S2, where a custom skill is installed on an Alexa enabled device. When the skill is used, the audio is sent to the Alexa service in the cloud for processing. The Alexa skill, set up by the developer in the Alexa Developer Console, processes the audio input and then textual data is sent to a backend web service, also set up by the developer, that hosts the backend conversational logic of the Alexa skill. We use a Lambda function on the Amazon Web Service (AWS) to host the web service and use a MariaDB database hosted on the Relational Database Service of the AWS to store the collected data. An overview of the system design is shown in Supplementary figure S3 and further details of the system architecture are provided in Supplementary Section S2.

*System configuration to improve response speeds*
Warm starts: A skill using a lambda function can be slow to start if the lambda function needs to be reprovisioned before it is ready. We use a CloudWatch timer to keep the lambda function “warm” so that response times are minimised.

Memory and CPU provisioning: We increased the lambda memory from 512MB to 1024MB which also increases the CPU power “proportionately”.

#### Supplementary section S3: Analytical sample

Of the 7 participants excluded with incorrect equipment, 4 were due to an equipment delivery error where the wrong equipment boxes were delivered to the wrong participants such that their web food diaries were mixed up and could not be used. The other 3 had home internet that was incompatible with the technology used in this study, namely use of a mini router to connect the EchoDot device to the participants home internet service.

#### Supplementary section S4: Summaries of responses to free-text questionnaire items

<u>Participation questionnaire</u>

*Other reason participant took part*

Two participants provided a response. One was interested in the application of new technologies to self-reported data, and the other wanted to see what the experience was like for a participant of a project using a novel technology for remote research.

*Other comments about your experience supplying dietary information via Alexa / Other things that happened when using Alexa*

32 participants provided additional comments. The comments were focused around three broad themes: 1) using the Alexa skill, 2) articulating or remembering/being able to provide information, and 3) and comments relating the study protocol.

Comments relating to using the Alexa skill (from 30 participants): 21 participants commented on the difficulty they had adding items using Alexa, such as difficulty providing Alexa with meals that had several components or non-English meals, and difficulties with inadvertent shutdown of the Alexa skill (e.g., due to a gap in speech). Of these 21 participants, 4 said this meant they simplified what they said so that Alexa could understand, and two said this meant they sometimes did not record whole events. Connection or speed issues were commented on by 12 participants and a further two mentioned times when the skill stopped working. Two participants commented that they would sometimes accidently add their information to the Alexa shopping list skill rather than the study custom skill. Two participants said they preferred writing the information compared with using Alexa while one said they preferred using Alexa. One participant noted the ‘now’ functionality to record the time did not work for them. 10 participants commented on the sentence structure required by Alexa being difficult, including not allowing pauses in speech, having to say ‘add item’ which some found inconvenient, and the general learning curve for using the skill. One participant commented that the effort needed meant they reduced the amount they snacked to avoid having to add an entry. Three participants noted they had difficulties using the skill generally, such as with inadvertent interruptions or skill shutdowns. One participant commented that they felt more connected using the web form compared with using the Alexa skill.

Comments relating to articulating or remembering/being able to provide information (from 11 participants): Nine participants commented that it could be difficult to articulate information to Alexa, for example, quantities and ingredients, with one participant who forgot to record quantities, another unsure on the amount of detail that was required, and another who was unsure whether different items in a meal should be in the same event. Three participants commented that they did not include water intake in their diaries, with one of these also noting they also did not add herbal teas as it does not have

[many] calories. One participant noted that they did not add diary information on one day due to illness, and another noted that they often did not record their events soon after they occurred.

Comments relating to the study protocol (from 9 participants): Five participants said that it was difficult when they ate or drank away from home, because their Alexa device was at home. Two participants made comments regarding the web form, one that Alexa would have been easier if they did not also need to use the web form, and the other that they did not fill in the web form which they suggested was because Alexa is easier. One participant commented that they hoped they would feel better about providing their food intake because they did not have to type it out, but that they did have to do that using the web form in this study. One participant noted that they did not know if Alexa had recorded what they told them (our study setup did not allow the participants to see the information they provided).

*Other reasons participant is not happy to use a voice-controlled system (e.g. Alexa) at home for research in the future?*
Five participants provided a response. Of which, four participants commented on the issues they encountered such as having to repeat information and having disruptions when using it. The remaining participant commented that using the wearable would be more convenient compared with using a home-based device.

*Other reasons participant is not happy to use a voice-controlled system (e.g. Alexa) on a wearable device such as a smart watch, for research*
One participant provided an additional reason, stating that it was because they needed to repeat information.

<u>Non-participation questionnaire</u>
*Other reasons given for not taking part, provided in non-participation questionnaire*
Of the 21 respondents who provided an additional reason, 15 of these said they did not receive or see the email inviting them to participate in the study. Other reasons given were not meeting the eligibility criteria, being busy or distracted with other things in their lives at the time the study took place, considering participating after recruitment had ended, 'lazyness', and the belief that Alexa only understands English foods.

*Other reasons respondent is not happy to use a voice-controlled system (e.g. Alexa) at home for research in the future?*
Of the 3 respondents who provided an additional reason, one cited privacy concerns and one talked about the effort required. The other talked about only want to pay for a device if it works well but as devices would be provided this individual has misunderstood.

#### Supplementary section S5: Further study strengths and limitations

<u>Further strengths</u>
We originally planned a study based at the University, and successfully adapted the study protocol to be 'contactless' so that it could be carried out during the pandemic.

<u>Further limitations</u>
We originally planned a study based at the University, and successfully adapted the study protocol to be 'contactless' so that it could be carried out during the pandemic.

While we provided an information sheet with the Alexa device to help participants to get started with Alexa, more comprehensive training may be useful, for example, providing a training video or asking them to try a test submission in the presence of a researcher who can advise them.

While asking participants to provide information via both Alexa and a web form was valuable, interactions with one of these approaches may have impacted their interaction and perceived feelings towards the other. Specifically, we asked participants to enter the information using Alexa first and then the web form, so the web form entry might be affected by their interaction with Alexa. For example, if the participant felt they could not enter much detail using Alexa, then they would likely then enter the same level of detail using the web form, but if this was reversed then we might be able to see how the information is reduced/changed due to the voice interface. Likewise, the participants experience using Alexa may have been impacted by having to also complete a web form.

We collected food and drink information via Alexa and the web form with no explicit link between them, asking the participants to enter the intake information via the web form after submitting this to Alexa. Contrary to our expectations, participants often did not enter the web entry directly after the Alexa entry and sometimes submitted them at completely different times. This meant it was difficult to identify the entries of these two approaches that corresponded to the same intake event. Many of the entries were excluded from our comparison of the submitted entries because we could not identify the counterpart entry entered with the other method.

#### SUPPLEMENTARY FIGURES

##### Supplementary figure S1: Participant flow diagram

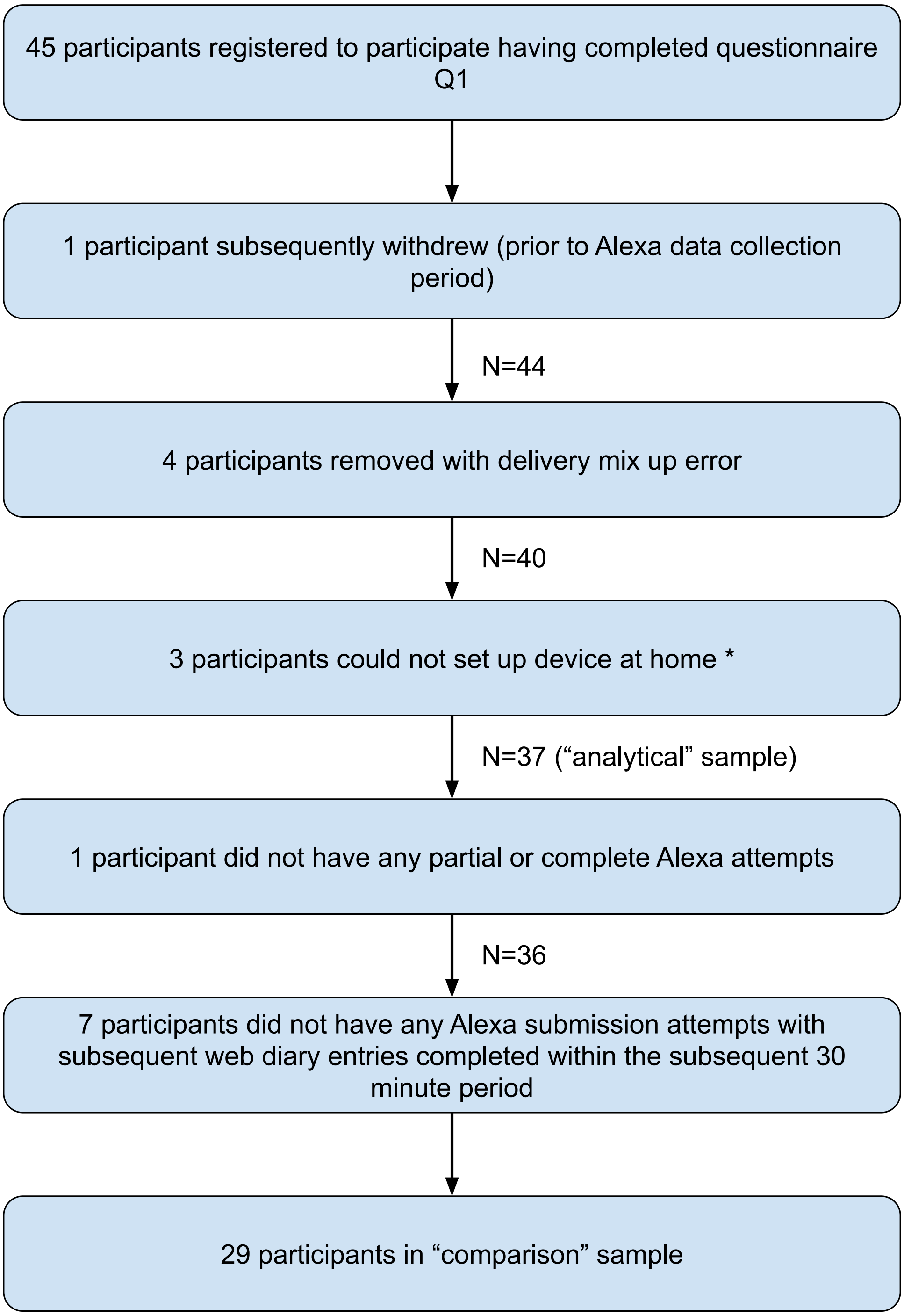

* This was due to the router-based set up of this study rather than due to the Alexa technology, see Supplementary section S3 for further details.

#### Supplementary figure S2: Infrastructure of custom Alexa skill

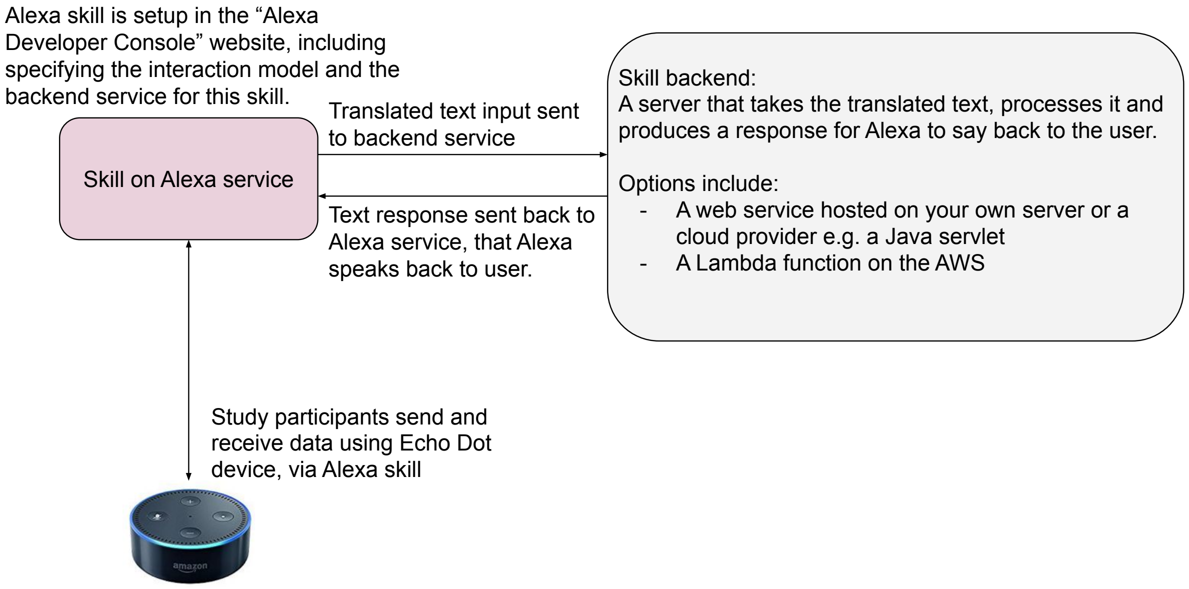

#### Supplementary figure S3: System architecture

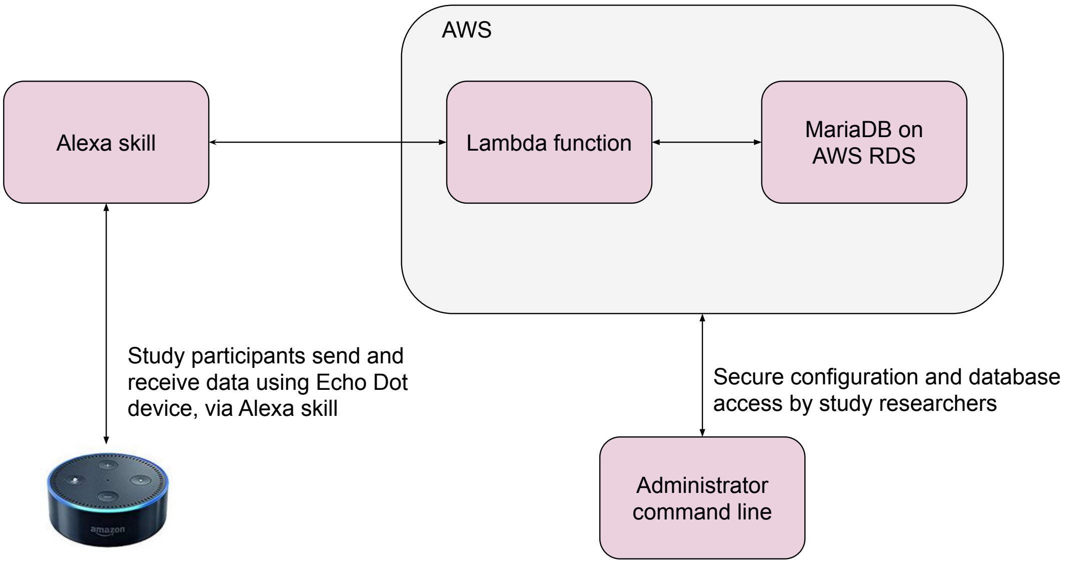

The Amazon Echo Dot device listens for the keyword 'Alexa' and then audio following this is sent to the Amazon web service. This web service parses the audio and uses our Alexa 'skill' to convert to text information. This textual data is then sent to our web service (hosted as an AWS lambda function), which stores the information in a MariaDB database on the AWS RDS.

#### Supplementary figure S4: User journey of food diary skill

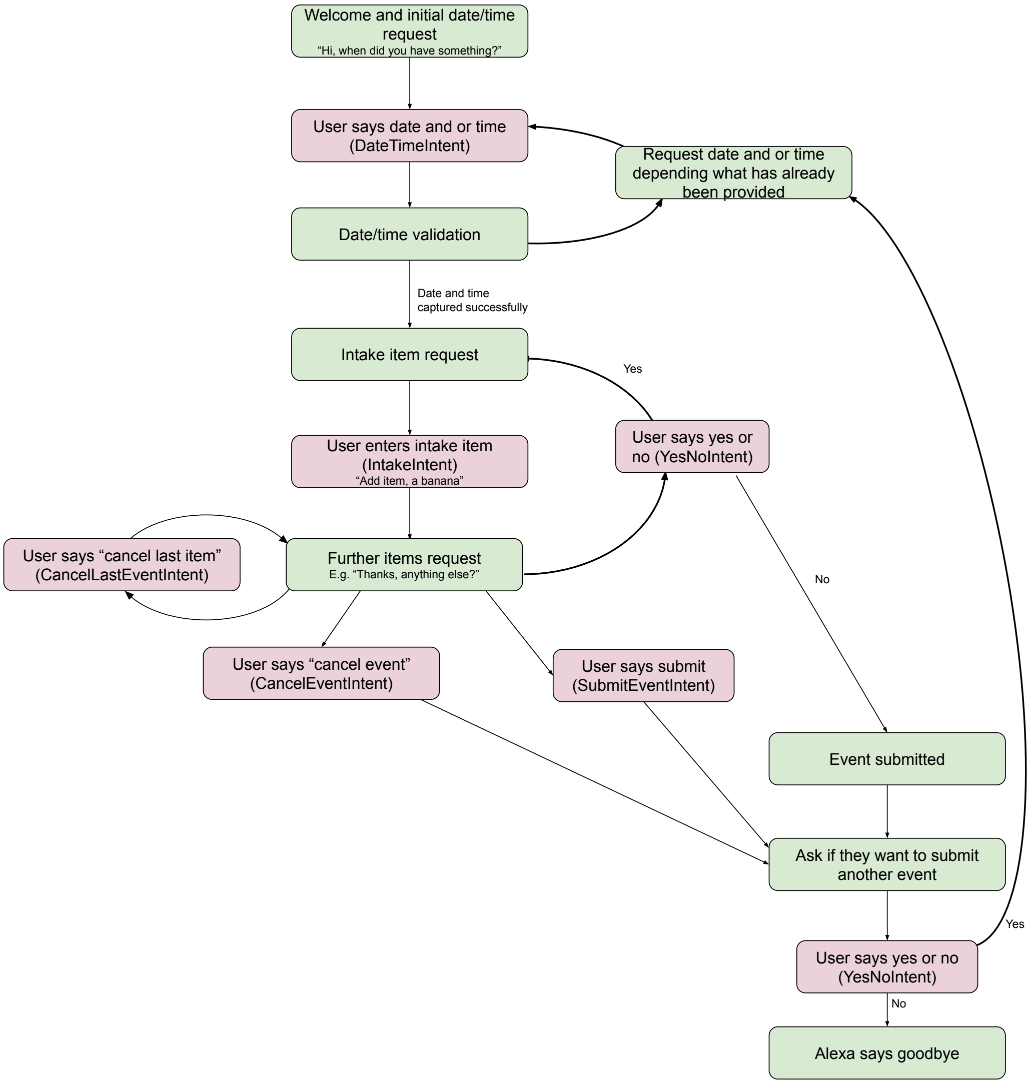

Supplementary figure S5: Food diary data processing approach

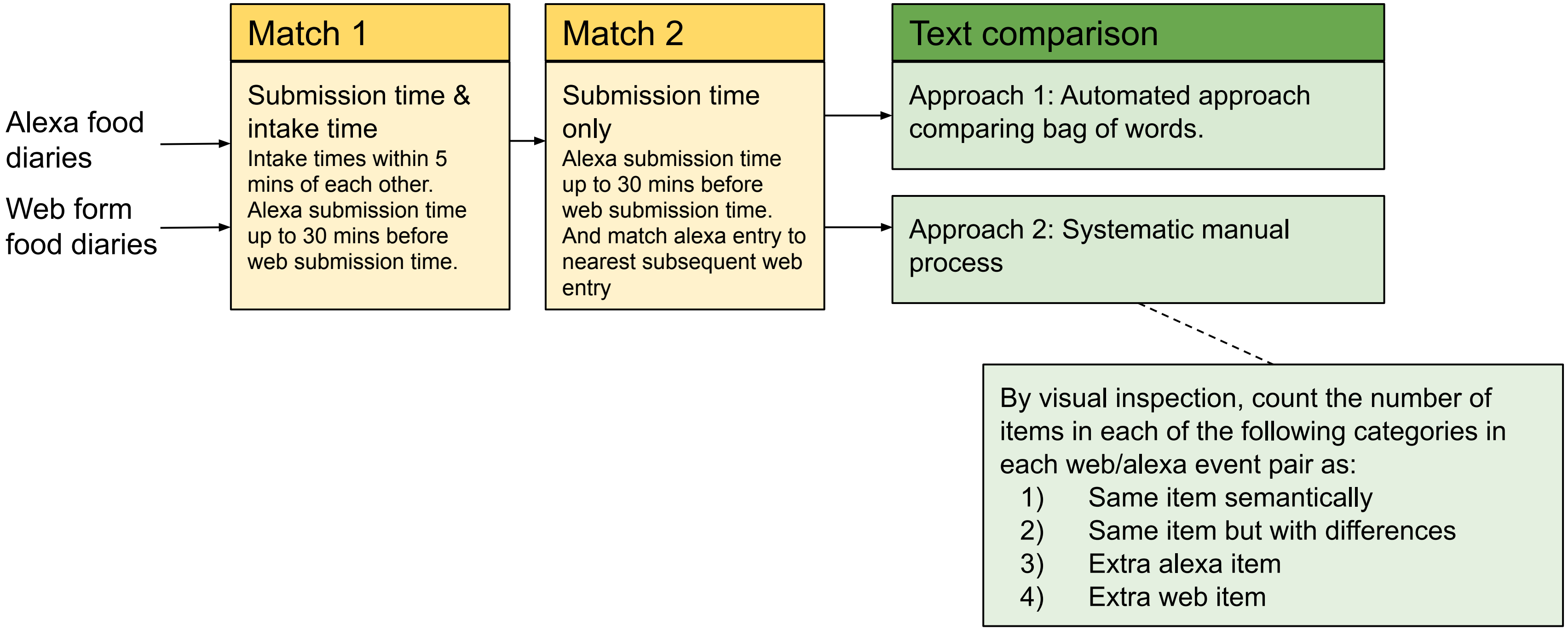

Supplementary figure S6: Summaries of the number of diary entries entered via the web form and Alexa

a) Number of web form entries versus Alexa entries

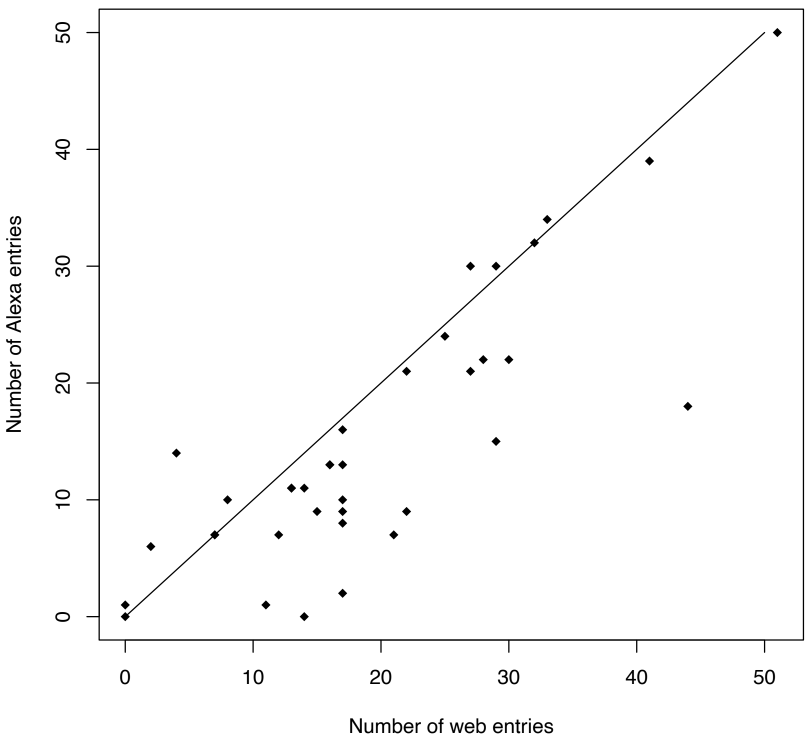

b) Number of partial Alexa entries

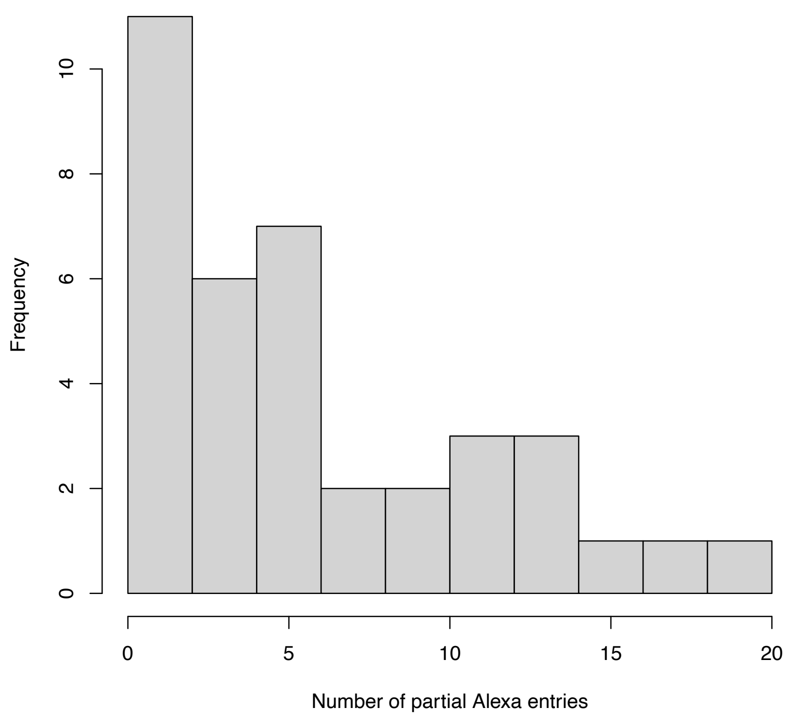

These summaries are of the main analytical sample, N = 37.

Supplementary figure S7: Comparison of systematic manual approach assessments of LACM versus independent assessors

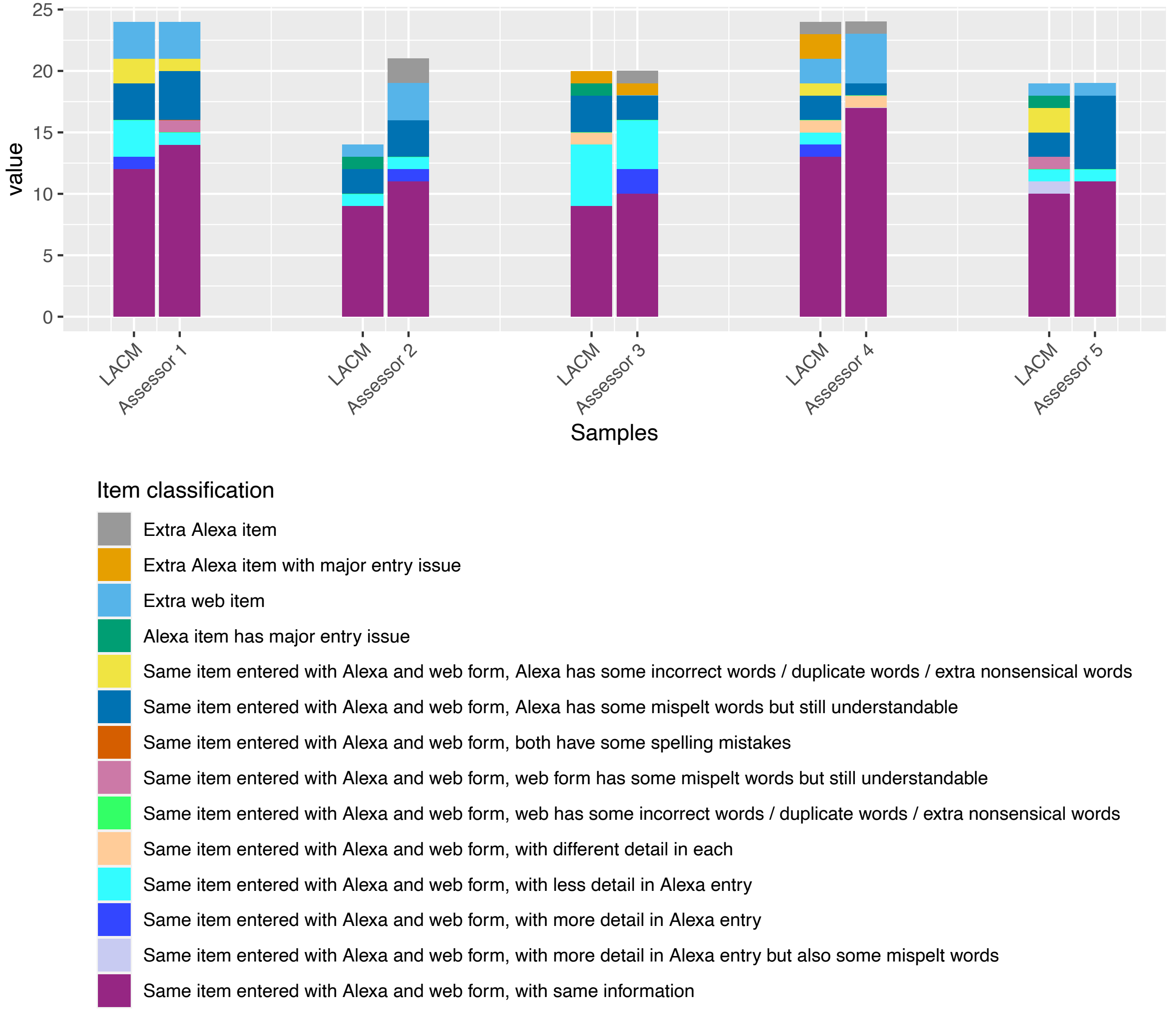

Assessor bars contain the summary of assessments for the set of examples randomly assigned to that assessor. Next to each are the assignments by LACM, for that same set of examples.

#### SUPPLEMENTARY TABLES

##### Supplementary table S1: Skill intents and example utterances

| Intent | Description | Example utterances |
|---|---|---|
| Intake event | Add food or drink item to intake event. | "Add item, a cheese and salad sandwich on brown bread"<br>"Yes add item, a cup of tea" |
| Date and time | Submit date and time of intake event. | "An hour ago"<br>"Ten thirty"<br>"Yesterday at 4pm" |
| Cancel event | Cancel the current intake event and start from the beginning. | "Cancel event"<br>"Start over" |
| Cancel last item | Remove the last intake item from the current intake event. | "Remove last item"<br>"Delete last item" |
| Yes No | Reply to a yes/no question during the conversational logic. For example, when Alexa asks the participant if they have other items or events to submit. See Figure 2 in main paper. | "Yes"<br>"No" |
| Submit event | Submit this intake event. | "Submit event"<br>"Submit" |

#### Supplementary table S2: Examples of manual text comparison

| **Web entry** | **Alexa entry** | **Result** |
|---|---|---|
| **Examples comparing individual items** | | |
| 1 x apple | One apple | This is the same item because the meaning is equivalent. Add 1 to *same_item* column. |
| A bowl of porridge | Some porridge | This is the same item but with extra info in the web entry. Add 1 to *same_item_alexa_less_detail* column. |
| 20g chocolate | 20 | Here there is no indication that it is referring to a different item, but it is not recognizable as this item. Add 1 to item_with_major_alexa_entry_issue column. |
| A small fruit bowl | A small football | The alexa item is not recognizable as fruit. Add 1 to item_with_major_alexa_entry_issue column. |
| Tomato pasta topped with cheese | Tomato pasta topped with cheddar | This is the same item but with different detail ('cheese' vs 'cheddar'). Add 1 to *same_item_alexa_different_detail* column. |
| A handful of cashew nuts | 15 cashew nuts | This is the same item but with different detail ('handful' vs a specific quantity). Add 1 to *same_item_alexa_different_detail* column. |
| Cup of tea | Cup of tea with milk | This is the same item but with extra info in the alexa entry (i.e. including that the tea was with milk). Add 1 to *same_item_alexa_more_detail* column. |
| A coffee with milk | A cough with milk | 'Coffee' is incorrectly recorded as 'cough' by Alexa. Add 1 to *same_item_alexa_entry_issue* column. |
| 2 x bagels with peanut butter | To bagels with peanut butter | The quantity is recorded by Alexa as 'to' rather than 'two'. Add 1 to *same_item_alexa_entry_issue* column. |
| **Example comparing whole intake event** | | |
| 20g chocolate<br>A coffee with milk<br>A bagel with peanut butter | A cough with milk<br>20 | Step 1: Note that there are 3 web items but only 2 Alexa items. Add this to the *total_web_items* and *total_alexa* items columns of the spreadsheet.<br><br>Step 2: Match the items that most likely correspond to each other:<br>"A coffee with milk" -> "A cough with milk"<br>Add 1 to the same_item_alexa_mispelt column.<br><br>"20g chocolate" -> "20"<br>While the Alexa entry is sparse, it has some similarity with the chocolate web item so this is the most likely corresponding item. Something has gone wrong entering this information with Alexa, so add 1 to the *item_with_major_alexa_entry_issue* column.<br><br>3. Identify the items that cannot be matched. In this case, it is the "A bagel with peanut butter" web entry. Add 1 to the *extra_web_item* column. |
