## Supplementary material for "“Alexa, I just ate a donut”: A pilot study collecting food and drink intake data with voice input": S1 file

Bristol Medical School
Dr Louise A C Millard


University of BRISTOL

**Study Information Sheet**

### Using Amazon's Alexa system to collect food and drink consumption information in epidemiology studies: a feasibility study

Thank you for expressing interest in this study. This information sheet will provide an outline of the aims of the study and describe what you will be asked to do as a participant in the study. We will also describe what will happen to the data that we collect from you. If you have any questions about any aspect of the study, please contact Dr Louise Millard, the study lead (contact details above).

#### What is the study about?

Epidemiology cohorts commonly collect information about the food and drink participants consume. Typically, participants are asked to complete a food frequency questionnaire or food diary, either on paper, through a web application (on a computer or smart phone) or (more recently) in some studies by photographing their food prior to eating. These approaches can require significant time and effort for participants, and participants may forget to complete the questionnaire/diary or take photos, or it may not convenient for them to do so.

The purpose of this study is to assess an alternative way to collect food diaries, using Amazon's Alexa system. Alexa is a voice-controlled system that can be installed in a room and used for a variety of tasks such as retrieving information, setting reminders and storing information provided by a user. In general, Alexa listens for the word 'Alexa', and after this is recognised it processes the subsequent request spoken by a user, to determine the correct action to take. For example, a user asking "Alexa, what is the time?" will be told the current time. Request processing is performed remotely on an Amazon server, and a blue light on the device indicates that Alexa is currently processing a verbal request. If Alexa is inadvertently activated, she will stop after a period of silence or can be stopped immediately using the command 'Alexa stop'.

#### Who is doing this study?

This study will be carried out by researchers at the MRC Integrative Epidemiology Unit at the University of Bristol. Dr Louise Millard is the lead researcher for this study.

#### Who is eligible to take part in this study?

Unfortunately, due to the small number of participants in this pilot study and risk of identification, those with speech impediments are not eligible to take part. You also need to live in a BS postcode to take part in this study.

#### What you will be asked to do

We will ask that you register for the study by emailing the study administrator (Shirley Jenkins,). You will be assigned a unique ID number that will be used to identify you. The administrator will store this ID alongside your name, email address and home address, and agree with you a 7-day period for you to use the Alexa device at home.

After registration you will be asked to complete a short initial questionnaire asking, for example, your age and sex, and also asking for your consent to participate in this study.

We will provide you with an Alexa-enabled device (an Echo Dot), delivered by courier to your home. We will ask you to plug this device in and provide us with information about what you eat and drink by telling this information to Alexa. Any time after you eat or drink we ask that you verbally tell Alexa the food and drink you have consumed. You can also submit food or drink from the day before, such as what you ate for dinner last night. After using Alexa, we also ask that you submit this information as written text on this web form, so that we can compare this information to the details collected using Alexa.

We ask that you submit your dietary information as many times as you are able, throughout the duration of this data collection exercise (you may make several submissions on the same day).

Specific instructions explaining how to submit your dietary information using Alexa will be provided with the device. After this 7-day period a courier will collect the device from your home.

We will then ask you to complete a second short questionnaire on your experiences during the study, and your thoughts and feelings on using voice-controlled systems in general. We will also invite those who have not participated so far to complete a questionnaire on their thoughts and feelings of using such technology.

#### How will your data be used and stored?

We will use the data collected to assess how accurately Alexa stores food diary data, by comparing the data collected using Alexa to that entered on the web form. We will assess how the accuracy varies within and between study participants, e.g. whether it varies by age and sex. We will also use your data to build predictive models, for example, to recognise whether words in descriptions of food or drink are describing the quantity (e.g. 'a **handful** of grapes').

The data collected via both Alexa and web questionnaires will be anonymous. We will ask you to select which age band you are in, rather than provide your specific age, so that risk of identification based on age and sex is minimised. The administrator will securely store your unique identifier, name, email address home address and the dates you use Alexa during the study. Only the unique identifier and Alexa usage dates will be provided to researchers so that we can link the Alexa data to your questionnaire responses. Your name, email address and home address will not be provided to study researchers.

In accordance with General Data Protection Regulations (GDPR), data submitted via the online questionnaires will be stored securely on a University of Bristol server, and will only be used by researchers working on this project.

We will use a dedicated Amazon account for this study, that will be linked to the Alexa device, so that any information collected by Alexa is linked to this Amazon account rather than your own. When your verbal food diary information is submitted using Alexa, data will be collected by the study researchers at University of Bristol, and will also be recorded and processed by Amazon. Only commands spoken after Alexa 'hears' the word 'Alexa' are recorded and processed by Amazon, and a blue light on the echo dot device will indicate this. The verbal recording will be processed by Amazon on a remote server including translating the verbal input to text, and then we store the translated text (in accordance with GDPR) in a secure database in the cloud (hosted by Amazon). After data collection is complete we will download the data to a University of Bristol server and delete the data in the cloud-based database. We (University of Bristol researchers) will not store the audio recordings submitted using Alexa – these are stored by Amazon and can be accessed by study researchers through the dedicated study Amazon account. After initial analyses we may access these recordings via the Amazon account to explore why Alexa might sometimes be inaccurate.

Amazon stores Alexa usage information such as the interactions performed (i.e. commands or information provided after the "Alexa" command), the users location, and some audio recordings of Alexa commands. Further details on the data that Amazon retains can be found in Section 1.3 of Alexa's Terms of Use. We are aware of these Terms of use so if you would like to discuss any aspect of these please contact Dr Millard (contact details at the top of this sheet). Amazon may use these data (audio or translated text) for purposes outside those of this study. The data stored by amazon will not be linked to identifying information (such as your name and address or your own amazon account) as all study data collected using Alexa will be linked to our dedicated Amazon account for this study.

While Alexa devices have 'drop-in' functionality that would allow communication between devices, this is controlled solely by PI Dr Millard and will be disabled throughout the duration of the study.

*While Alexa is set up in your home environment, it is possible that other data may be inadvertently collected if Alexa 'hears' the word 'Alexa'.*

#### What are the benefits of participating in this study?

You would be contributing to new knowledge that may improve the quality of information that studies collect on diet and the burden placed on study participants to collect these data.

#### What if I do not want to take part, or want to stop participating?

It is your choice whether you take part or not. If you decide to take part and then want to stop being in the study you can ask us to remove all your existing data that has been stored by the study researchers. As described above, information submitted using Alexa may also be collected by Amazon, and it is not possible to ask Amazon to delete your data.

#### How has the risk of coronavirus been minimised during study participation?

The Alexa device will be cleaned with disinfectant wipes between each participant's turn with the device. The device will be delivered to and collected from your home via a courier, in a contact-free manner. To minimise risk to other participants, before the device is delivered to you we will ask you to confirm that you do not have coronavirus symptoms and are not currently self-isolating. While it is up to you whether you take part in this study, we advise that those at high risk of becoming seriously ill from coronavirus do not take part.

#### Who has reviewed this study?

This study has been reviewed by the Faculty of Health Science Research Ethics Committee.

#### Who can I contact about this study?

For further information about the study, please contact Dr Louise Millard using the contact details at the top of this sheet.

Should you wish to make a formal complaint about any aspect of this study or its implementation, please write to:.
