## Supplementary material for "“Alexa, I just ate a donut”: A pilot study collecting food and drink intake data with voice input": S2 file

Millard et al.

**SUPPLEMENTARY FILE S2**

### Q1: Online questionnaire

Please answer the following questions to the best of your knowledge.

Please provide your unique participant identifier *: ______________________________

#### Section A: Consent

| | YES | NO |
|---|---|---|
| **HAVE YOU:** | | |
| • been given information explaining the study? | ☐ | ☐ |
| • had an opportunity to ask questions and discuss this study? | ☐ | ☐ |
| • received satisfactory answers to all questions you asked? | ☐ | ☐ |
| • received enough information about the study for you to make a decision about your participation? | ☐ | ☐ |
| **DO YOU UNDERSTAND:** | | |
| • that you are free to withdraw from the study at any time and without having to give a reason for withdrawing? | ☐ | ☐ |
| • that it is possible to request that your data is destroyed by contacting the study administrator | ☐ | ☐ |

I hereby fully and freely consent to my participation in this study.

I understand the nature and purpose of this study. This has been communicated to me on the information sheet accompanying this form.
I understand and acknowledge that the study is designed to promote scientific knowledge.
I understand that, although we will delete voice recordings from the study's dedicated Amazon account at the end of the study, Amazon may still store Alexa usage information such as the interactions performed (i.e. commands or information provided after the "Alexa" command) and the device location, and it is not possible to ask Amazon to delete these data.

Date: [auto populated]

☐ Please check this box to confirm your consent.

**Having participated in this study**

I agree to the University of Bristol keeping and processing the data I provide for this study.
I understand that the study researchers will use these data only for the purpose(s) set out in the information sheet, and my consent is conditional upon the University complying with its duties and obligations under the Data Protection Act and the General Data Protection Regulation (GDPR).

Date: [auto populated]

☐ Please check this box to confirm your consent.

### Section B: Participant questionnaire

**A. Please select your sex *:**

☐ Male

☐ Female

☐ Other

☐ Prefer not to answer

**B. Please select your age band *:**

☐ <20

☐ 20-29

☐ 30-39

☐ 40-49

☐ 50-59

☐ 60+

☐ Prefer not to answer

**C. Do you consider yourself to have a strong UK regional accent? ***

Yes
A little
No
Prefer not to answer

**D. Do you consider yourself to have a strong accent due to English being a second language? ***

Yes
A little
No
Prefer not to answer

E. Do you currently have a voice-controlled device (e.g Alexa) at home?

Yes and I use it
Yes but it’s used by others, not me
No

### Q2: Participation questionnaire

This questionnaire is for those who **did** participate in the Alexa pilot study.

Please answer the following questions to the best of your knowledge.

** Required fields*

**Section A: Taking part in this study**

1. Please provide your unique participant identifier *: ______________________________

2. Please tell us the reason(s) you **did** take part (please select all that apply):
   a) Interested in nutritional research.
   b) I wanted to support research.
   c) I wanted to support others in the IEU.
   d) I feel I should be a research participant when asked, as I benefit from other participants.
   e) I wanted to try a voice controlled system.
   f) I wanted to try Alexa.
   g) I wanted to see how well Alexa could interact with me.
   h) I wanted to see how well Alexa could record my dietary information.
   i) Other (please write any other reasons) ……………………………

3. Please tell us about your experience supplying your dietary information

Score each 1 to 5 where 1 = completely agree; 2 = agree somewhat; 3= neutral; 4= disagree somewhat; 5= disagree completely.

a. I was able to accurately tell Alexa what I ate or drank.
b. I was able to estimate accurate quantities describing how much I ate.
c. I chose not to record particular snacks/meals (e.g. because it was unhealthy).
d. Sometimes, I chose to be selective with the truth.
e. I remembered to submit my food/drink information.
f. Other comments ………………………………………….

4. Please tell us how often the following things happened when using Alexa in this study
Score each 1 to 5 where 1 = Never; 2 = Rarely; 3= occasionally; 4= Often; 5= Always.

a. Alexa interjected when I hadn't finished telling her **when** I ate or drank.
b. Alexa interjected when I hadn't finished telling her **what** I ate or drank.
c. Other comments ………………………………………….

5. How convenient or inconvenient did you find providing information using Alexa?
   a. Very inconvenient
   b. Somewhat inconvenient
   c. OK
   d. Somewhat convenient

e. Very convenient

6. How enjoyable or unenjoyable did you find providing information using Alexa?
   a. Very unenjoyable
   b. Somewhat unenjoyable
   c. OK
   d. Somewhat enjoyable
   e. Very enjoyable

7. How efficient or inefficient did you find providing information using Alexa?
   a. Very inefficient
   b. Somewhat inefficient
   c. OK
   d. Somewhat efficient
   e. Very efficient

8. How easy or hard did you find providing information using Alexa?

   Very easy
   Somewhat easy
   OK
   Somewhat hard
   Very hard
   I could not use Alexa at all.

Other comments ………………………………………….

10. Did you have any concerns about taking part in this study (please select all that apply)?

   a) Data privacy concerns around Amazon collecting information on my diet.
   b) Data privacy concerns around researchers collecting information on my diet.
   c) Concerns that Alexa will inadvertently listen to other conversations.
   d) other (please write all other reasons) ...................

**Section B: Previous experience recording a diet diary**

9a. Have you ever used another method to collect diet information? If so, please enter them here starting with those you have use the most used (if you have used more than three then give the three you have used the most often).

9b.i. Which method? *Please specify details (e.g. which website) in the details box below.*

   a) A website
   b) An app on your phone
   c) An app on your tablet device (e.g. iPad)
   d) A traditional diary (on paper or a document on the computer)
   e) Other

Details of method used (e.g. which website or app you used): ……………………….

9c.ii. How convenient or inconvenient was Alexa to use compared with this method?

a. Much less convenient
b. A little less convenient
c. About the same
d. A little more convenient
e. Much more convenient

9c.iii. How enjoyable or unenjoyable was Alexa to use compared with this method?

a. Much less enjoyable
b. A little less enjoyable
c. About the same
d. A little more enjoyable
e. Much more enjoyable

9c.iv. How efficient or inefficient was Alexa to use compared with this method?

a. Much less efficient
b. A little less efficient
c. About the same
d. A little more efficient
e. Much more efficient

Section C: Using voice-controlled devices for research in the future

12. Would you be happy to use a voice-controlled system (e.g. Alexa) at home for future research studies? *If you don't have one then assume it would be provided by the research group.*

Yes
No
Not sure

If <u>NO or NOT SURE:</u> Please tell us the reason(s) you would <u>not</u> (be sure you would) be happy to use a voice-controlled system at home for future research studies (please select all that apply):

a) Data privacy concerns around researchers collecting information about me.
b) Data privacy concerns around Amazon collecting information about me.
c) Concerns that the system will inadvertently listen to other conversations, which may then be accessed by researchers.
d) Concerns that the system will inadvertently listen to other conversations, which may then be used by the company that owns it (e.g. Amazon).
e) other ...................

13. Would you be happy to use a voice-controlled system (e.g. Alexa) on a wearable device such as a smart watch, for research? *If you don't have one then assume it would be provided by the research group.*

Yes
No

Q2: Participation questionnaire

Not sure

If <u>NO or NOT SURE:</u> Please tell us the reason(s) you would <u>not</u> (be sure you would) be happy to use a voice-controlled system on a wearable device for research (please select all that apply):

a) Data privacy concerns around researchers collecting information about me.

b) Data privacy concerns around Amazon collecting information about me.

c) Concerns that the system will inadvertently listen to other conversations, which may then be accessed by researchers.

d) Concerns that the system will inadvertently listen to other conversations, which may then be used by Amazon.

e) I wouldn't like the idea of wearing a device all the time because of how it looks and / or feels.

f) other ...................

### Q3: Non-participation questionnaire

This questionnaire is for those who **did not** participated in the Alexa pilot study.

#### Section A: Consent

| | YES | NO |
|---|---|---|
| • Have you been given information explaining about the study? | ☐ | ☐ |
| • Have you had an opportunity to ask questions and discuss this study? | ☐ | ☐ |
| • Have you received satisfactory answers to all questions you asked? | ☐ | ☐ |
| • Have you received enough information about the study for you to make a decision about your participation? | ☐ | ☐ |
| Do you understand that it is not possible to request that your data is destroyed as it is collected anonymously | ☐ | ☐ |

I hereby fully and freely consent to my participation in this study.

I understand the nature and purpose of this study. This has been communicated to me on the information sheet accompanying this form.
I understand and acknowledge that the study is designed to promote scientific knowledge and that the University of Bristol will use the data I provide for no purpose other than research.
I understand the data I provide will be anonymous. No link will be made between my name or other identifying information and my study data.

Date: [auto populated]

☐ Please check this box to confirm your consent.

**Having participated in this study**
I agree to the University of Bristol keeping and processing the data I provide for this study.
I understand that the study researchers will use these data only for the purpose(s) set out in the information sheet, and my consent is conditional upon the University complying with its duties and obligations under the Data Protection Act and the General Data Protection Regulation (GDPR).

Date: [auto populated]

☐ Please check this box to confirm your consent.

### Section B: About you

1. Please select your sex: *

    Male
    Female
    Other
    Prefer not to answer

2. Please select your age band: *

    <20
    20-29
    30-39
    40-49
    50-59
    >=60
    Prefer not to answer

3. Please tell us the reason(s) you **<u>did not</u>** take part (please select all that apply):

    a) Not available during the study session times.
    b) Data privacy concerns around Amazon collecting information on my diet.
    c) Data privacy concerns around researchers collecting information on my diet.
    d) Concerns that Alexa will inadvertently listen to other conversations.
    e) I don’t eat or drink during my working hours
    g) other (Please write all other reasons) ...................

4. Do you currently have a voice-controlled device (e.g Alexa) at home?

    Yes and I use it
    Yes but it’s used by others, not me
    No

5. In the future, would you be happy to use a voice-controlled system (e.g. Alexa) at home for research?
*If you don’t have one then assume it would be provided by the research group.*

    Yes
    No
    Not sure

If <u>NO or NOT SURE:</u> Please tell us the reason(s) you would <u>not</u> (be sure you would) be happy to use a voice-controlled system at home for research (please select all that apply):

    a) Data privacy concerns around researchers collecting information about me.
    b) Data privacy concerns around Amazon collecting information about me.
    c) Concerns that Alexa will inadvertently listen to other conversations, which may then be accessed by researchers.

d) Concerns that the system will inadvertently listen to other conversations, which may then be used by the company that owns it (e.g. Amazon).

e) other ...................

13. Would you be happy to use a voice-controlled system (e.g. Alexa) on a wearable device such as a smart watch, for research? *If you don't have one then assume it would be provided by the research group.*

Yes

No

Not sure

If NO or NOT SURE: Please tell us the reason(s) you would not (be sure you would) be happy to use a voice-controlled system on a wearable device for research (please select all that apply):

a) Data privacy concerns around researchers collecting information about me.

b) Data privacy concerns around Amazon collecting information about me.

c) Concerns that the system will inadvertently listen to other conversations, which may then be accessed by researchers.

d) Concerns that the system will inadvertently listen to other conversations, which may then be used by Amazon.

e) I wouldn't like the idea of wearing a device all the time because of how it looks and / or feels.

f) other ...................

### Q4: Food and drink diary intake event form

*Form fields marked with a * are required.*

Please select the date and time of this event.

Time of event: *

Participant ID: *

What did you eat and/or drink? *
*Tell us about the food and drink you had. How much did you have?*

Please rate this Alexa interaction. *
*Terrible* *Ideal*
0 ◯ 1 ◯ 2 ◯ 3 ◯ 4 ◯ 5 ◯

Please provide any comments on your interaction with Alexa.
