## Supplementary material for "“Alexa, I just ate a donut”: A pilot study collecting food and drink intake data with voice input": S3 file

** QR CODE PLACED HERE **

Scan to access web forms

### Alexa Pilot Study: Participant Guide

*If you have any problems taking part, please contact Louise Millard,, or the study administrator Shirley Jenkins,.*

#### EQUIPMENT PACKAGE

The equipment package contains 2 devices:

*Echo Dot device with power cable and plug*

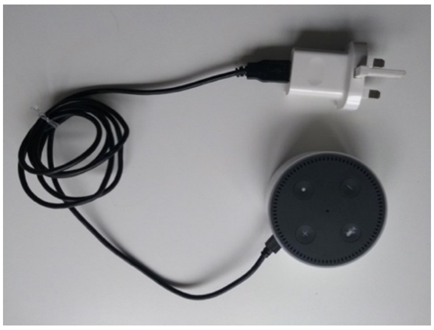

*Mini router with ethernet cable, power cable and plug*

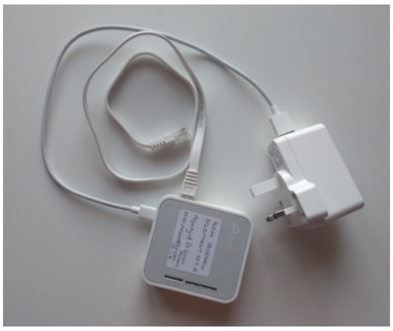

*For the purposes of this study, we are providing an Echo Dot that is already linked to our study Amazon account. In order to connect this device to the internet at your home, it is also already set up to connect to our mini router, which can be plugged into your home internet router.*

*An extension cable is also included for use if needed.

#### ECHO DOT DEVICE AND ROUTER SETUP

1. Plug the router plug into a power socket and the router ethernet cable into an ethernet socket on your home's internet router.

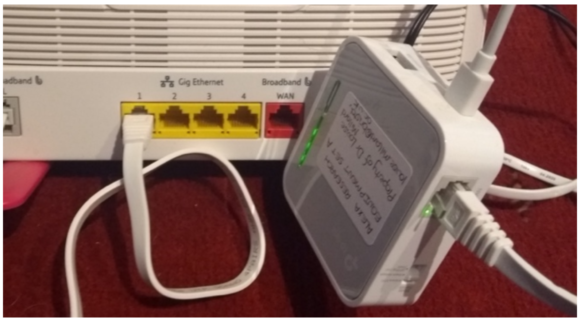

2. Plug the Echo Dot into the power supply, anywhere in your home. For example, you could put it in the kitchen or near where you usually eat. If you already have an Alexa, put this one in a different room.

3. Wait for the light around the device to turn blue and then stop.

4. Test the Echo Dot by saying 'Alexa hello'.

#### SUBMITTING YOUR FOOD AND DRINK INFORMATION

After you have had something to eat or drink, we would like you to submit your food and drink information to Alexa <u>first</u>, and then submit it on the web form (you will have received an email with the web address for this).

Our Alexa 'skill' collects information on *what* you ate and drank, *when*, and *how much*.

It is important you also enter the food/drink/time on the web form, so that we can compare this to the information entered using Alexa.

Open the food diary by saying: **"Alexa, open food diary".** This opens our custom food diary skill, which is like an app. You can then converse with Alexa to tell her what you have eaten and drunk. Only information you submit within this skill is sent to our study database.

**You will interact with our skill by adding intake events – any time when you have eaten and/or drunk something. Each event may have many items.** For example, if for lunch you had a houmous and salad sandwich, with crisps, and a cup of tea, we would like you to add the three items separately with a quantity for each:

<u>Intake event at 1:10pm:</u>

<u>Item 1</u>: one houmous and salad sandwich on brown bread
<u>Item 2</u>: one packet of salt and vinegar crisps
<u>Item 3</u>: one cup of tea with milk

Your conversation might go something like this:

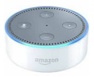

Alexa open food diary

Hi, when did you have something?

Today at one ten pm

You can also say:
'just now'
'this evening at one ten'
'yesterday evening at ten past one'

… don't forget to indicate if it is morning or afternoon!

Thanks, please add items

Start with 'add item' to add an item

Add item, a houmous and salad sandwich on brown bread

Great, anything else?

Add item, one packet of salt and vinegar crisps

OK anything else?

Add item, a small cup of tea with milk

See **portion size guide** for examples ways you can describe the amounts.

Thanks, anything else?

No

OK do you have other events to submit?

No

OK thanks, goodbye!

If you find that too arduous you can be more concise, for example:

Add item, a houmous and salad sandwich, a packet of crisps and a cup of tea

**Other things you can say**

Clear the *current event* and start over by saying:

Cancel event

Clear the *last item in the current event* by saying:

Cancel item

#### TALKING TO ALEXA

You should be able to talk fairly naturally to Alexa, but some tips are:

- *Do* speak clearly
- *Do* project towards the Echo Dot device
- *Don't* interrupt Alexa – when she's speaking she can't hear you!
  **A *blue ring with a stationary cyan element on the device indicates she is listening*** (see other status indicators on the next page).

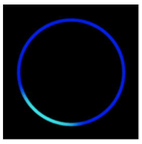

- *Do* tell Alexa whether the event happened in the morning or afternoon "e.g. 7 O'clock in the evening" or "5 PM".
- *Don't* worry! If it goes wrong please try again and report any challenges in the online food form.

*If at some point you want to temporarily stop Alexa from 'listening' you can use the microphone one/off button on the top of the device. A red light indicates the microphone is off.*

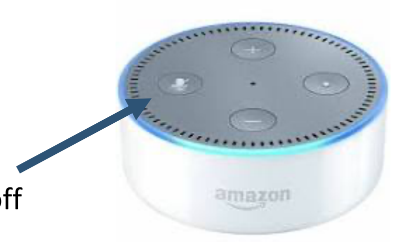


Microphone on / off button

### Portion Size suggested descriptors

Here are some example descriptions to help you describe how much you eat or drink.

##### EATING EXAMPLES

**1 individual pot of** yoghurt

**2** vegetarian sausages

**1 medium** jacket potato

**A small bowl** of porridge made with water

**A handful of** grapes

**Two fists** of mashed potato

**About 2 tablespoons of** yoghurt

**Half a can of** baked beans

**A matchbox amount** of cheddar cheese

**About one ladle** of gravy

##### DRINKING EXAMPLES

**A small glass** of red wine

**One medium glass of** milk

**A large mug** of hot chocolate, half water half milk

**Half a pint** of weak orange squash

**A pint of** corona beer

### Alexa status indicators

Alexa displays colour patterns to let you know its status. Here are the main ones you might see:

**Blue ring with *spinning* cyan element**

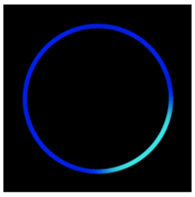

Device is starting up. You should see this when you first plug the Alexa in. Then the lights stop then Alexa is ready to use.

**Blue ring with *stationary* cyan element**

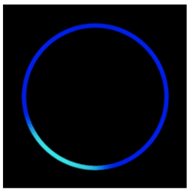

Alexa is listening to what you say.

**Orange, spinning**

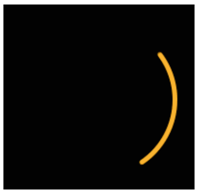

Device is trying to connect to the internet.

**White**

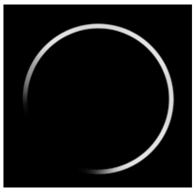

Indicates volume levels, displayed when you are adjusting the volume.

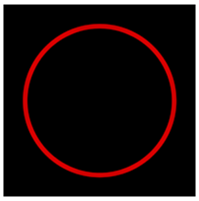

**Red full circle**
Microphone button has been pressed such that microphone is off and Alexa isn't listening. You can press the microphone button again to turn the microphone back on.

Each status indicator has a different light movement. To see these go to:

https://www.amazon.com/gp/help/customer/display.html?nodeId=GKLDRFT7FP4FZE56

Scan to view status indicators
