## Supplementary material for "“Alexa, I just ate a donut”: A pilot study collecting food and drink intake data with voice input": S4 file

Millard et al.

### SUPPLEMENTARY FILE S4

### Email 1: participant study invitation

Subject: Participants needed! Using Amazon Alexa for health research – home-based study

Email Contents

Dear All,

* Apologies for the cross-posting *

You are invited to take part in a research study using cutting edge technology – Amazon's Alexa device.

Alexa is a voice-controlled system that can be used for a variety of tasks such as retrieving information, setting reminders and storing information provided by a user.

We are looking to find out whether Alexa can be used to collect information about people's food and drink consumption in epidemiology studies.

To thank you for taking part you will receive a **£30 Love2Shop voucher** that you can spend in a wide range of shops**.**

You need to live in a BS postcode to take part in this study.

Unfortunately, those with speech impediments are not eligible to take part, due to the small number of participants in this pilot study and risk of identification.

Please see the attached participant information sheet for further information.

If you have any queries or issues taking part in this study, please do not hesitate to contact myself or Dr Louise Millard <> directly.

Kind regards,

Shirley Jenkins, study administrator

### Email 2a: participant invitation reply 1 – choosing dates

Dear [PARTICIPANT_NAME],

Thank you for agreeing to participate in this study.

Please complete an initial questionnaire here:
*https://sscmredcap.bris.ac.uk/redcap/surveys/?s=KRFH3HCNK9&participant=1&q1_pid=XXX*

Please let me know your address, and also advise which of the following dates would be suitable for you to receive and use the Alexa, to collect details of your food and drink intake:

XXX-XXX
XXX-XXX
XXX-XXX

The Alexa will the delivered to you between 12pm – 2pm on day 1 and collected between 12pm – 2pm on day 7. Could you please confirm if these times would be suitable for delivery and collection?

Also, it would be helpful for the courier to have a contact telephone number for you, if you would be happy to provide this.

Kind regards,

Shirley Jenkins,
Study administrator

### Email 2b: Participation invitation reply 2 – confirmation

Dear [PARTICIPANT_NAME],

Thank you, I have booked you in for the period between [XXX-XXX], and will send a reminder to you the day before.

Please ensure you are able to be at home between 12-2pm on the start day to receive the equipment from the courier.

In the meantime, please feel free to contact me if you have any questions about the study.

Kind regards,

Shirley Jenkins,
Study administrator

### Email 3: Reminder for start of Alexa usage period

Subject: Reminder: Your Alexa pilot study data collection period is starting [tomorrow]

Dear [PARTICIPANT_NAME],

Thanks for agreeing to participate in this study.

Your Alexa period starts [tomorrow (XX September)], when a courier will drop off the equipment between [12-2pm]. You will need to be at home during these times to receive the equipment.

Please let us know if you have COVID-19 symptoms or are self-isolating as we will need to reschedule your Alexa period.

Here is a link to your online survey page including the food diary form. We ask that you complete this straight away, after you submit your food and drink via Alexa: https://sscmredcap.bris.ac.uk/redcap/surveys/?sq=[XXXXXXXXX]

This is important so that we can compare the data you enter via Alexa with a written diary.

The equipment box will be collected on [XX September between 12-2pm]. Please note that we ask that you setup the device as soon as possible after you receive it (preferably the same day); it will not be possible to extend your Alexa data collection period.

We hope you enjoy taking part. Please get in touch if you have any issues or questions during this pilot.

Kind regards,

Shirley Jenkins,
Study administrator

### Email 4: Reminder for end of Alexa usage period

Subject: Reminder: Your Alexa pilot study data collection period is ending [tomorrow]

Dear [PARTICIPANT_NAME],

We hope you have been getting on well with Alexa.

Your Alexa period ends [tomorrow (XX May)]. A courier will collect the equipment between [12-2pm].

**Please ensure the equipment box contains the following:**

1) **Mini router and two cables (1 ethernet cable and 1 power cable)**
2) **Echo dot (Alexa device) and power cable**
3) **Extension power cable**

Thanks for taking part!

Kind regards,

Shirley Jenkins,
Study administrator

### Email 5: Post-participation questionnaire

Subject: Using Amazon Alexa for health research, post-participation questionnaire

Dear [PARTICIPANT_NAME],

Thanks for taking part in the Alexa pilot study.

Please complete a final questionnaire that should take no longer than 10 minutes to complete, and can be access via the following link:

*https://sscmredcap.bris.ac.uk/redcap/surveys/?s=KRFH3HCNK9&participant=1&q1_pid=[PID]*

If you have any queries or issues taking part in this study, please do not hesitate to contact myself or Dr Louise Millard <> directly.

Many thanks,

Shirley Jenkins
Study administrator

### Email 6: Invitation for non-participation questionnaire

Subject: Using Amazon Alexa for health research, follow-up questionnaire for those who **did not** participate

Dear Oakfield House / IEU staff member,

We recently emailed inviting you to take part in our research study – collecting food and drink information with the Amazon Alexa voice-controlled system. This study aims to assess whether Alexa can be used to collect food and drink information from participants in epidemiology studies.

The food diary data collection stage of this study is now complete, thanks very much to those who took part.

We now invite **all staff members who did not participate in the main study,** to complete a questionnaire on your experiences using voice-controlled systems and your feelings around using such technology.

The questionnaire should take no longer than 10 minutes to complete.

Unfortunately, those with speech impediments are not eligible to take part, due to the small number of participants in this pilot study and risk of identification.

Please see the attached participant information sheet for further information.

If, after reading the participant information sheet you decide to take part, please click on the following link to complete the questionnaire:

https://sscmredcap.bris.ac.uk/redcap/surveys/?s=KRFH3HCNK9&participant=0

If you have any queries or issues taking part in this study, please do not hesitate to contact myself <> or Dr Louise Millard <> directly.

Many thanks,

Shirley Jenkins, study administrator
