## Supplementary material for "“Alexa, I just ate a donut”: A pilot study collecting food and drink intake data with voice input": S5 file

### Manual comparison of web vs Alexa entries

You are asked to review each row of the spreadsheet you have been sent. Each row has a web entry containing a number of food/drink items entered via a web form, and an Alexa entry containing a number of food/drink items entered using the Amazon Alexa system. Each entry contains one or more intake *items*, and these are separated by two vertical bars " || ".

Use your own judgement when you think there is ambiguity in what you should do.

The evaluation process we would like you to undertake has two steps.

#### Step 1: Decide if the entries correspond to the same intake event

We ask you to first compare the items in these entries, and complete the *different_eating_occurrence* column, with a value of either 0 or 1:

- Give this column value 1 if the entries are clearly corresponding to different intake events i.e. they both contain understandable content about food or drink with no overlap in the food or drink that was consumed. For example, "A plate of chips and an apple" vs "a bowl of spaghetti bolognaise".
- Otherwise, give this column a value 0. I.e. if this is clearly corresponding to the same intake event, or if you are not sure.

Only those assigned as 0 in the *different_eating_occurrence* column are taken forward to the next step to compare the text content.

#### Step 2: Compare items in each Alexa and web entry pair

Do this step for all rows assigned *different_eating_occurrence*=0 in step one.

**Step 2.1: Decide which items in the web entry correspond to which items in the Alexa entry.**

- Match the items in the web entries with the items in the Alexa entry. If the Alexa and web entries have different splits of items then use the most granular version for that item. For example, if the web entry has 1 item "salad and tomato pasta", and the Alexa entry has two items: "salad" and "tomato pasta", treat these as two items in both the Alexa and web entries.
- Fill in the *total_alexa_items* and *total_web_items columns with the total numbers entered with each approach* (yellow highlighted columns).
- Fill in the *extra_alexa_item* and *extra_web_item* columns with the total numbers of items in each approach, that do not have a corresponding item with the other approach (grey highlighted columns).

Here are some examples:

**Example A:**

| Web entry | Alexa entry | Result |
|---|---|---|
| 20g chocolate \|\| A coffee with milk \|\| A bagel with peanut butter | A cough with milk \|\| 20 | The web entry has three items and the Alexa entry has two items.<br><br>I've matched them in this way:<br><br>"20g chocolate" / "20"<br>"A coffee with milk" / "A cough with milk"<br>"A bagel with peanut butter" / MISSING<br><br>So, the values I give are:<br>*total_web_items=3* and *total_alexa_*items=2 |

| | | |
|---|---|---|
| | | *extra_alexa_item=0 and extra_web_item=1* |

**Example B:**

| Web entry | Alexa entry | Result |
|---|---|---|
| A jacket potato with cheese \|\|<br>salad with lettuce, tomatoes and cucumber | jacket potato\|\|<br>cheese \|\|<br>salad with lettuce and tomatoes \|\|<br>cucumber\|\|<br>mayonnaise | The web entry has two items and the Alexa entry has five items.<br><br>This example is more complicated because the web and Alexa entries have different granularity (e.g A jacket potato with cheese is one item in the web entry but two items in the Alexa entry) . In order to know how many items in total there are, we first need to identify item groups, which are the items in each entry that correspond but have different granularity.<br><br>You need to use your judgment to say when different items in each entry should be grouped.<br><br>Group 1:<br>The web item “A jacket potato with cheese” matches with Alexa items: “jacket potato” “cheese”<br>- the highest granularity is with the Alexa entry which has 2 items so we split the web entry items accordingly:<br><br>“A jacket potato” / “jacket potato”<br>“with cheese” / “cheese”<br><br>Group 2:<br>“salad with lettuce, tomatoes and cucumber” matches with “salad with lettuce and tomatoes”, “cucumber”<br>- the split with the most granularity is the Alexa entry with 2 items:<br><br>“salad with lettuce, tomatoes” / “salad with lettuce and tomatoes”<br>“and cucumber” / “cucumber”<br><br>Group 3:<br>The Alexa item “mayonnaise” does not have a matching item in the web entry, so it is placed in its own group.<br><br>MISSING / “mayonnaise”<br><br>There are two items in group 1, two items in group 2, and one item in group 3. The item in group 3 only occurs in the Alexa entry so the values we give are:<br>*total_web_items=4* and *total_alexa_*items=5<br>*extra_alexa_item=1 and extra_web_item=0* |

**Example C:**

| Web entry | Alexa entry | Result |
|---|---|---|
| Spaghetti bolognaise with broccoli. A can of diet coke. | Spaghetti bolognaise \|\|<br>broccoli | Sometimes a participant did not split the web entry into items. In this scenario, split into item groups using your own judgment. In this example, I will put the coke in a different group as (using my judgment) most people would consider this a different intake item. |

| | | |
|---|---|---|
| | | Group 1:<br>"Spaghetti bolognaise" / "Spaghetti bolognaise"<br>"with broccoli". / "Broccoli"<br><br>Group 2:<br>"A can of diet coke." / MISSING<br><br>There are two items in group 1, and 1 item in group 2 (that is only in the web entry). So, the values I give are:<br>*total_web_items=3* and *total_alexa_*items=2<br>*extra_alexa_item=0 and extra_web_item=1* |

*Note: A repeated item is counted as an extra item (i.e., it doesn't matter that it is the same thing). For example, if the web entry has one item 'spaghetti bolognaise' and the Alexa entry has two items 'spaghetti bolognaise' and 'spaghetti bolognaise', then there is one matching item and the Alexa entry has one extra item (i.e. extra_alexa_item=1 and same_item=1).*

**Step 2.2: Compare the item pairs**
Classify each item pairings into one of the following categories.

Please read each category to understand them before attempting the task. The categories are disjoint (i.e. an item can be put in only one category), such that the totals across these categories should equal the totals identified in step 2.1 above. If you feel an items belongs in multiple categories, choose the one you think matters most.

**same_item**: The web and Alexa entries are equivalent and neither has extra detail compared with the other.

| Web entry | Alexa entry | Result |
|---|---|---|
| 1 x apple | One apple | This is the same item because the meaning is equivalent. Increment *same_item* column by one. |

**same_item_different_detail:** You believe the Alexa item to be the same as the web item but it contains different detail, probably because the participants submitted different information via the two methods.

| Web entry | Alexa entry | Result |
|---|---|---|
| Tomato pasta topped with cheese | Tomato pasta topped with cheddar | This is the same item but with different detail ('cheese' vs 'cheddar'). Add 1 to *same_item_different_detail* column*.* |
| A handful of cashew nuts | 15 cashew nuts | This is the same item but with different detail ('handful' vs a specific quantity). Add 1 to *same_item_different_detail* column*.* |

**same_item_alexa_less_detail:** You believe the Alexa item to be the same as the web item but it contains less detail.

| Web entry | Alexa entry | Result |
|---|---|---|
| A bowl of porridge | Some porridge | This is the same item but with extra info in the web entry (i.e. a specific bowl quantity versus 'some'). Add 1 to *same_item_alexa_less_detail* column*.* |

**same_item_alexa_more_detail:** You believe the Alexa item to be the same as the web item but it contains more detail.

| Web entry | Alexa entry | Result |
|---|---|---|
| Cup of tea | Cup of tea with milk | This is the same item but with extra info in the Alexa entry (i.e. including that the tea was with milk). Add 1 to *same_item_alexa_more_detail* column. |

**same_item_alexa_more_detail_alexa_misspelt:** Alexa entry has a miss-spelling and more detail.

**same_item_alexa_misspelt:** You believe the items to be the same, but the Alexa entry has some incorrect words, either misspelling, homophone (sounds the same but spelt differently) or a completely different word. Don't worry about American English vs UK English spellings – treat these as the same word.

| Web entry | Alexa entry | Result |
|---|---|---|
| A coffee with milk | A cough with milk | 'Coffee' is incorrectly recorded as 'cough' by Alexa. Add 1 to *same_item_alexa_misspelt* column. |
| 2 x bagels with peanut butter | To bagels with peanut butter | The quantity is recorded by Alexa as 'to' rather than 'two'. Add 1 to *same_item_alexa_misspelt* column. |

**same_item_web_misspelt**: equivalent to same_item_alexa_misspelt.

**same_item_both_misspelt:** both the Alexa and web entries have a miss-spelling.

| Web entry | Alexa entry | Result |
|---|---|---|
| Two quorne chicken pieces | To quorn chicken pieces | The correct spelling is 'quorn', and Alexa has heard 'to' rather than 'two'. Add 1 to *same_item_both_misspelt* column. |

**extra_alexa_item_with_major_entry_issue:** There is an Alexa item that cannot be matched with a web item, for example, it might be a partial entry that was not entered in its entirety.

| Web entry | Alexa entry | Result |
|---|---|---|
| Three satsumas | 2 || 3 satsumas | There is an extra item '2' with a major issue (i.e. it does not contain any food or drink). Add 1 to *extra_alexa_item_with_major_entry_issue* column (and add 1 to *same_item* column for the 3 satsumas). |
| An apple | A tree || 1 apple | There is an extra item 'a tree' with a major issue (i.e. it does not contain any food or drink). Add 1 to *extra_alexa_item_with_major_entry_issue* column (and add 1 to *same_item* column for the apple). |

**item_with_alexa_entry_issue:** Same item, with Alexa entry issue, where the consumed item is still identifiable.

| Web entry | Alexa entry | Result |
|---|---|---|
| bowl of yoghurt | Ball of yoghurt | This is the same item but Alexa has heard 'bowl' as 'ball'. The main essence of the item (yoghurt) is still identifiable. Add 1 to *item_with_alexa_entry_issue* column. |

**item_with_web_entry_issue**: Equivalent to *item_with_alexa_entry_issue*.

**item_with_major_alexa_entry_issue**: A food or drink item is not recognisable in the Alexa item, but it has a corresponding web entry.

| **Web entry** | **Alexa entry** | **Result** |
|---|---|---|
| 20g chocolate | 20 | Here there is no indication that it is referring to a different item, but it is not recognizable as this item. Add 1 to *item_with_major_alexa_entry_issue* column. |
| A small fruit bowl | A small football | The Alexa item is not recognizable as fruit. Add 1 to *item_with_major_alexa_entry_issue* column. |

The following examples demonstrate how to match the items in the web and Alexa entries, for examples A, B and C used in step 2.1.

**Example A:**

| **Web entry** | **Alexa entry** | **Result** |
|---|---|---|
| 20g chocolate \|\| A coffee with milk \|\| A bagel with peanut butter | A cough with milk \|\| 20 | The web entry has three items and the Alexa entry has two items.<br><br>I've matched them in this way:<br><br>"20g chocolate" / "20"<br>"A coffee with milk" / "A cough with milk"<br>"A bagel with peanut butter" / MISSING<br><br>So, the values I give are:<br>*total_web_items=3* and *total_alexa*_items=2<br>*extra_alexa_item=0 and extra_web_item=1*<br><br>Step 2: Match the items that most likely correspond to each other:<br>"A coffee with milk" -> "A cough with milk"<br>Add 1 to the same_item_alexa_misspelt column.<br><br>"20g chocolate" -> "20"<br>While the Alexa entry is sparse, it has some similarity with the chocolate web item so this is the most likely corresponding item. Something has gone wrong entering this information with Alexa, so add 1 to the *item_with_major_alexa_entry_issue* column. |

**Example B:**

| **Web entry** | **Alexa entry** | **Result** |
|---|---|---|
| A jacket potato with cheese \|\|<br>salad with lettuce, tomatoes and cucumber | jacket potato\|\|<br>cheese \|\|<br>salad with lettuce and tomatoes \|\|<br>cucumber\|\|<br>mayonnaise | The web entry has two items and the Alexa entry has five items.<br><br>This example is more complicated because the web and Alexa entries have different granularity (e.g A jacket potato with cheese is one item in the web entry but two items in the alexa entry) . In order to know how many items in total there are, we first need to identify item groups, which are the items in each entry that correspond but have different granularity.<br><br>You need to use your judgment to say when different items in each entry should be grouped.<br><br>Group 1:<br>The web item "A jacket potato with cheese" matches with Alexa items: "jacket potato" "cheese"<br>- the highest granularity is with the Alexa entry which has 2 items so we split the web entry items accordingly:<br><br>"A jacket potato" / "jacket potato"<br>"with cheese" / "cheese"<br><br>Both items ('jacket potato' and 'cheese') are provided in both the Alexa and web items, so same_item=2.<br><br>Group 2:<br>"salad with lettuce, tomatoes and cucumber" matches with "salad with lettuce and tomatoes", "cucumber"<br>- the split with the most granularity is the Alexa entry with 2 items:<br><br>"salad with lettuce, tomatoes" / "salad with lettuce and tomatoes"<br>"and cucumber" / "cucumber"<br><br>Both items ('salad with lettuce and tomatoes' and 'cucumber') are provided in both the Alexa and web items, so same_item=2.<br><br>Group 3:<br>The Alexa item "mayonnaise" does not have a matching item in the web entry, so it is placed in its own group.<br><br>MISSING / "mayonnaise"<br><br>This is not a paired item (with an Alexa and web entry) so step 2 does not apply.<br><br>There are two items in group 1, two items in group 2, and one item in group 3. The item in group 3 only occurs in the alexa entry so the values we give are:<br>*total_web_items=4* and *total_alexa_*items=5<br>*extra_alexa_item=1 and extra_web_item=0* |

| | | |
|---|---|---|
| | | We add up the numbers in each category, across item groups. In this case only the same_item category was used, so same_item = 2+2=4. |

**Example C:**

| **Web entry** | **Alexa entry** | **Result** |
|---|---|---|
| Spaghetti bolognaise with broccoli. A can of diet coke. | Spaghetti bolognaise \|\| broccoli | Sometimes the web entry may not be split into items. In this scenario, split into item groups using your own judgment. In this example, it will put the coke in a different group as (using my judgment) most people would consider this a different intake item.<br><br>Group 1:<br>"Spaghetti bolognaise" / "Spaghetti bolognaise"<br>"with broccoli". / "Broccoli"<br><br>Both items are provided in both the Alexa and web items, so same_item=2.<br><br>Group 2:<br>"A can of diet coke." / MISSING<br>This is not a paired item (with an Alexa and web entry) so step 2 does not apply.<br><br>There are two items in group 1, and 1 item in group 2 (that is only in the web entry). So, the values I give are:<br>*total_web_items=3* and *total_alexa_*items=2<br>*extra_alexa_item=0 and extra_web_item=1*<br><br>We add up the numbers in each category, across item groups. In this case only the same_item category was used in one group, so same_item = 2. |

**Step 2.3: Checking the totals add up**

Review the green columns to check the 'Alexa totals match' and 'Web totals match' columns contain only TRUE values. If they contain FALSE, then we ask that you review the 'Calculated Alexa total' and 'Calculated web total' columns, that add up all the counts you entered in the white columns, for the Alexa and web items, respectively.

We ask you to:

- Compare the 'Calculated Alexa total' column with the 'total_alexa_items' column to see the discrepancy, and correct the column counts so that these have the same value.
- Compare the 'Calculated web total' column with the 'total_web_items' column to see the discrepancy, and correct the column counts so that these have the same value.
